# Association of reproductive and gender-related characteristics with cardiovascular risk factors of women in India: an analysis of nationally representative data

**DOI:** 10.1101/2025.05.02.25326891

**Authors:** Vinita Subramanya, Shakira Suglia, K.M. Venkat Narayan, Suryakant Yadav, Robin A. Richardson, Alvaro Alonso, Shivani A. Patel

## Abstract

**Introduction:** Women have unique characteristics based on their biological sex and gender that could contribute to their cardiovascular risk. This paper investigates the association between reproductive and gender-related characteristics on cardiovascular risk factors (CVRF) among women in India.

**Methods:** This cross-sectional analysis included 54,200 women aged 18-49 years from the National Family Health Survey-5, restricted to participants interviewed for the state or domestic violence module. Associations of reproductive characteristics (age at first birth, use of contraception, the total number of children born, pregnancy loss) and gender-related characteristics (age at first union, access to financial resources, owning property, currently working, experience of intimate partner violence) with CVRF (diabetes, hypertension, overweight or obesity) were estimated with Poisson regression. Analyses stratified by socioeconomic status, measured by education, were also performed.

**Results:** Women who had their first childbirth before the age of 20 years had a higher prevalence of all assessed CVRF across socioeconomic groups. Pregnancy loss was associated with a higher prevalence of being overweight or obese [prevalence ratio (PR) (95% CI): 1.16 (1.09, 1.23)], while an early age at first union (< 20 years) was linked to a greater prevalence of hypertension [PR (95% CI): 1.09 (1.02, 1.17)] that persisted across socioeconomic groups. Intimate partner violence was associated with a higher prevalence of hypertension in women with a primary school education or lower [PR (95% CI): 1.15 (1.05, 1.25)].

**Conclusion:** The association of reproductive and gender-related characteristics with CVRF across the socioeconomic spectrum underscores the importance of integrated consideration of biological and social determinants to evaluate cardiovascular risk in women.

## Introduction

Although cardiovascular disease is the leading cause of death in women, women remain underrepresented in cardiovascular research. Moreover, health research among women has primarily focused on issues related to pregnancy and the postpartum period. Cardiovascular disease in women of reproductive age is uniquely shaped by both biological sex and gender-related factors. Sex—a biological construct determined by reproductive characteristics— influences the distribution of cardiovascular risk factors (CVRF) such as hypertension, diabetes, and obesity as well as cardiovascular outcomes.(1, 2, 3, 4, 5, 6, 7, 8) Gender—a multidimensional social construct shaped by an individual’s body, identity, and self-expression within a societal framework—(9) has been less well studied in relation to cardiovascular disease (CVD). While sex and gender are distinct concepts, they overlap because the societal construction of gender roles often incorporates biological sex characteristics.(6, 10, 11, 12) Understanding cardiovascular risk in women requires examining biological and social factors and their intersection.

With respect to reproductive characteristics unique to women, sex hormones such as estrogen and progesterone play a major role. While these hormones are typically cardioprotective and integral to reproduction, they can increase cardiovascular risk when present at non- physiological levels.(13, 14, 15) During reproduction, pregnancy-induced hemodynamic changes and hormonal fluctuations(16, 17) can contribute to this increased risk. Reproductive characteristics, such as age at menarche and menopause, parity, short birth intervals, and pregnancy outcomes, also influence the development of CVD in women.(18, 19, 20, 21, 22, 23, 24, 25)

With respect to gender, multiple direct and indirect influences on cardiovascular risk are supported by the literature.(6) Gender roles, a set of culturally and socially determined traits, influence health and lifestyle choices.(26, 27, 28, 29, 30, 31) Women often engage in sedentary behavior and experience unique stressors related to caregiving, work-family life, gender discrimination, finances, and experiences of violence and abuse, contributing to chronic stress.(6) Prolonged stress and inflammation can increase cardiovascular disease risk through multiple pathways, including elevated blood pressure, lipids, and blood glucose.(6) Patriarchal power dynamics favor men’s access to opportunities and resources, limiting women’s financial access, asset ownership, decision-making, and autonomy, which have a detrimental effect on health and act as a barrier to healthcare access and utilization.(26, 27, 28, 29, 32, 33)

India is a particularly rich setting to investigate the intersection of sex and gender in relation to CVD. Reproductive histories in India differ from those observed in high-income country settings, and strong social structures influence gender norms regarding health behaviors.(34) Disparities in socioeconomic development have further complicated the relationship between gender, social structures, and cardiovascular health. Prior work in India indicates higher CVRF prevalence among the socioeconomically advantaged but higher mortality among the disadvantaged.(35, 36, 37) Women disproportionately face health disparities driven by high inequality and poverty.(38, 39) This, along with shifting gender roles and related changes in lifestyles and health behaviors may impact CVD risk but have not yet been studied.(40, 41)

Focusing on a single aspect of the biological or social components of women’s unique risks constrains our understanding of health inequalities arising from multiple intersecting processes. Therefore, this study adopted a woman-focused conceptual framework to clarify the role of reproductive and gender-related characteristics on CVRFs across educational attainment. The objectives were to evaluate the cross-sectional associations between reproductive and gender-related characteristics with CVRFs and ii) examine whether observed associations between reproductive and gender-related factors differed by educational attainment.

## Methods

### Data sources and study population

The National Family Health Survey (NFHS), conducted by the International Institute for Population Sciences under the aegis of the Ministry of Health and Family Welfare, is a nationally representative, comprehensive, multi-round cross-sectional survey of households.(42) NFHS-5 (2019–21), the fifth and most recent round, included data from 724,115 women from 636,699 households recruited through a stratified two-stage sampling approach.(42) The sampling methodology has been described previously.(42) An additional state or domestic violence module with information on gender-related and socioeconomic characteristics was administered in a random sub-sample of 15% of participants, where one woman per eligible household was included.(42) This study was restricted to married women aged 18-49 years from the domestic violence module (N=72,320). The analytic sample (**Figure 1**) comprised 54,200 ever-married women with complete information for study covariates.

**Figure 1:**
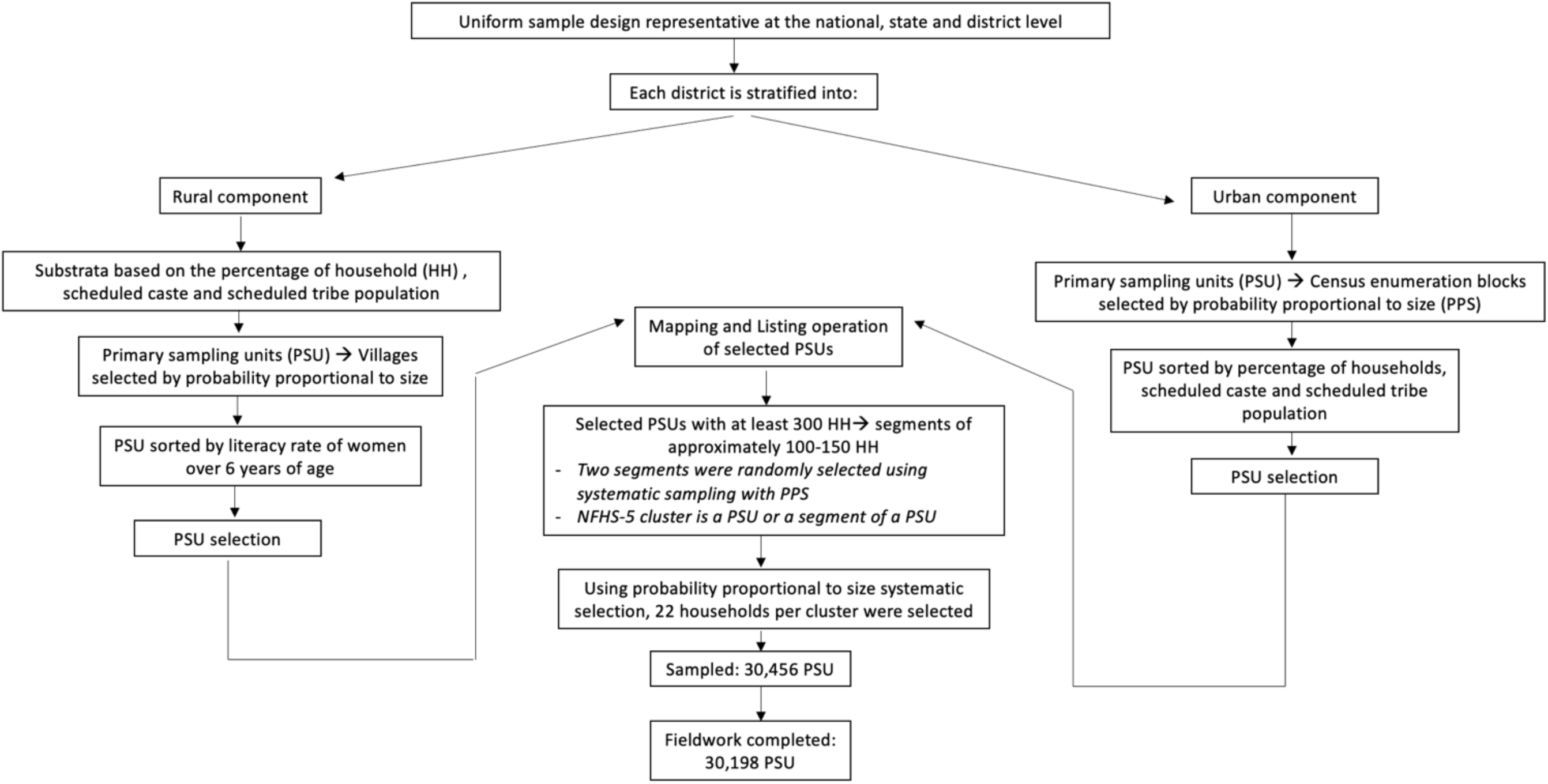
Sampling strategy used in National Family Health Survey-5

**Figure 2:**
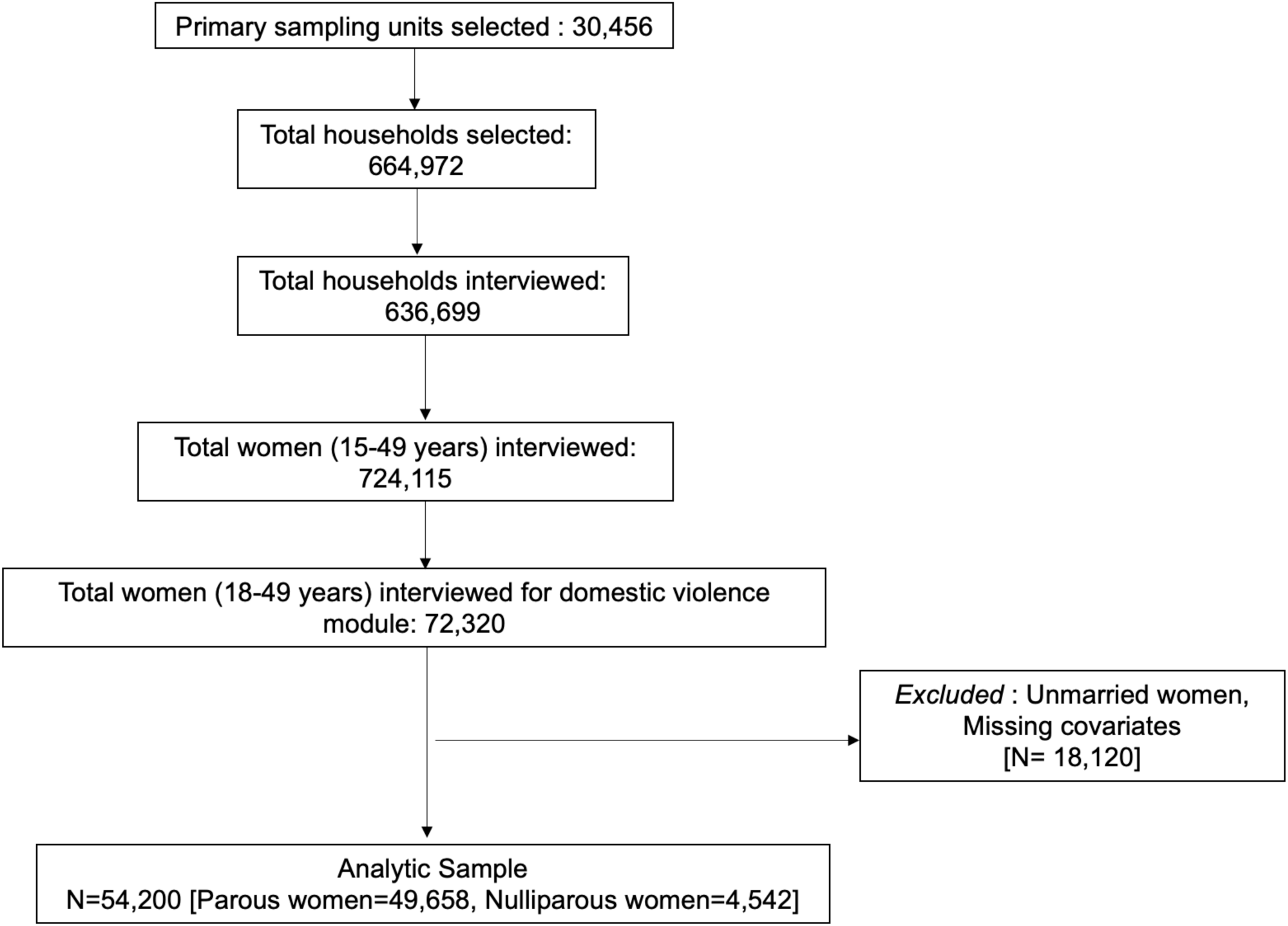
Participant flow diagram

### Study measures

The study utilized information from three instruments: a household survey collecting sociodemographic and socioeconomic details; a women’s survey assessing reproductive, marital, and sexual history, women’s empowerment, and domestic violence; and a biomarker assessment measuring anthropometry, blood pressure, and random blood glucose.

### 1. Cardiovascular risk factors

Diabetes was defined by the presence of any of the following: self-reported diabetes, use of glucose-lowering medication, a diagnosis of high blood glucose levels on at least two separate occasions, or a random blood glucose level exceeding 200 mg/dL. Hypertension was defined as a systolic blood pressure (SBP) of ≥140 mm Hg or a diastolic blood pressure (DBP) of ≥90 mm Hg or using blood pressure lowering medication or a diagnosis of high blood pressure on two separate occasions. These cut-offs were based on the Indian Guidelines for Hypertension- IV.(43) Height and weight were used to calculate the body mass index. Overweight and obese status were defined using a body mass index cutoff of >25 kg/m^2^.

### 2. Reproductive characteristics

Reproductive characteristics were self-reported and included use of contraception, the total number of children born, age at first birth (among those reporting any birth), and pregnancy loss. Contraceptive usage was categorized as (1) not using any method, (2) use of a modern method (sterilization, injectables, intrauterine devices, oral contraceptive pills, implants, barrier methods such as condoms and diaphragm, foam/jelly, standard days method, lactational amenorrhea method, and emergency contraceptive pills), and (3) use of a traditional method (including rhythm, withdrawal, and other traditional methods).(42) Pregnancy loss or having ever had a pregnancy not resulting in a live birth included information on spontaneous abortions, medical termination of pregnancy, and stillbirth.(42)

### 3. Gender-related characteristics

Gender-related indicators included those suggestive of a woman’s autonomy over their life choices for the improvement of their social, political, economic, and health status, such as age at first union (restricted to those ever reporting a union), access to financial resources such as owning and operating a bank account, owning property as well as, the experience of intimate partner violence (IPV). Information on IPV included any experience of physical, sexual, or emotional IPV and compared to having no experience of violence.(42, 44)

### 4. Socioeconomic and demographic variables

Socioeconomic status refers to an individual’s position within a stratified social structure, commonly determined by income, wealth, education, and occupation.(45, 46) In this study, educational attainment—a key indicator of socioeconomic status—was categorized into three levels: primary education or less, completed secondary education, and education beyond the secondary level. The current age of participants was analyzed as a continuous variable.

Religion was categorized as Hindu, Muslim, and ‘Others’ due to sparse categories. Social caste was self-reported as belonging to a scheduled caste or tribe or other backward class. Place of residence was classified as urban or rural.

### Statistical analysis

Data was described using mean (standard deviation) or percentage for continuous and categorical variables, as appropriate. In the primary analysis, associations of reproductive and gender-related characteristics with CVRFs were evaluated using Poisson regression models with robust variance estimation to directly estimate prevalence ratios.(47) Models were adjusted for sociodemographics and state of residence. For analyses evaluating reproductive characteristics, we stratified the sample by parity, as parous women who have experienced childbirth differ in their risk profile compared to nulliparous women who have not experienced childbirth. We estimated models with multiplicative interactions between the primary exposures of interest (reproductive and gender-related characteristics) and educational attainment to evaluate whether associations differed by educational attainment. Analyses applied state module survey weights and were conducted using Stata SE (version 18).

## Results

### Distribution of socioeconomic, reproductive, and gender-related characteristics

Less than 10% of the study population comprised nulliparous women (N=4,542). Among parous women, the mean (SD) age of the women in the study was 35.0 (8.1) years (**Table 1**). About 70% of participants lived in rural parts of India. About 12% of women had completed secondary or higher education, and 37% were employed. Forty-four percent of parous women had their first childbirth between 15-19 years of age. Examination of gender-related characteristics showed that over 95% of all women were married for the first time by age 30 (**Table 1**), and 30% experienced some form of intimate partner violence. The overall prevalence of hypertension was 23%, diabetes was 6%, and overweight or obesity was 29%. Nulliparous women were younger, with a mean (SD) age of 26.2 (8.0). About 30% of them had completed secondary school or higher. The prevalence of CVRFs was lower in nulliparous women as compared to parous women.

**Table 1:**
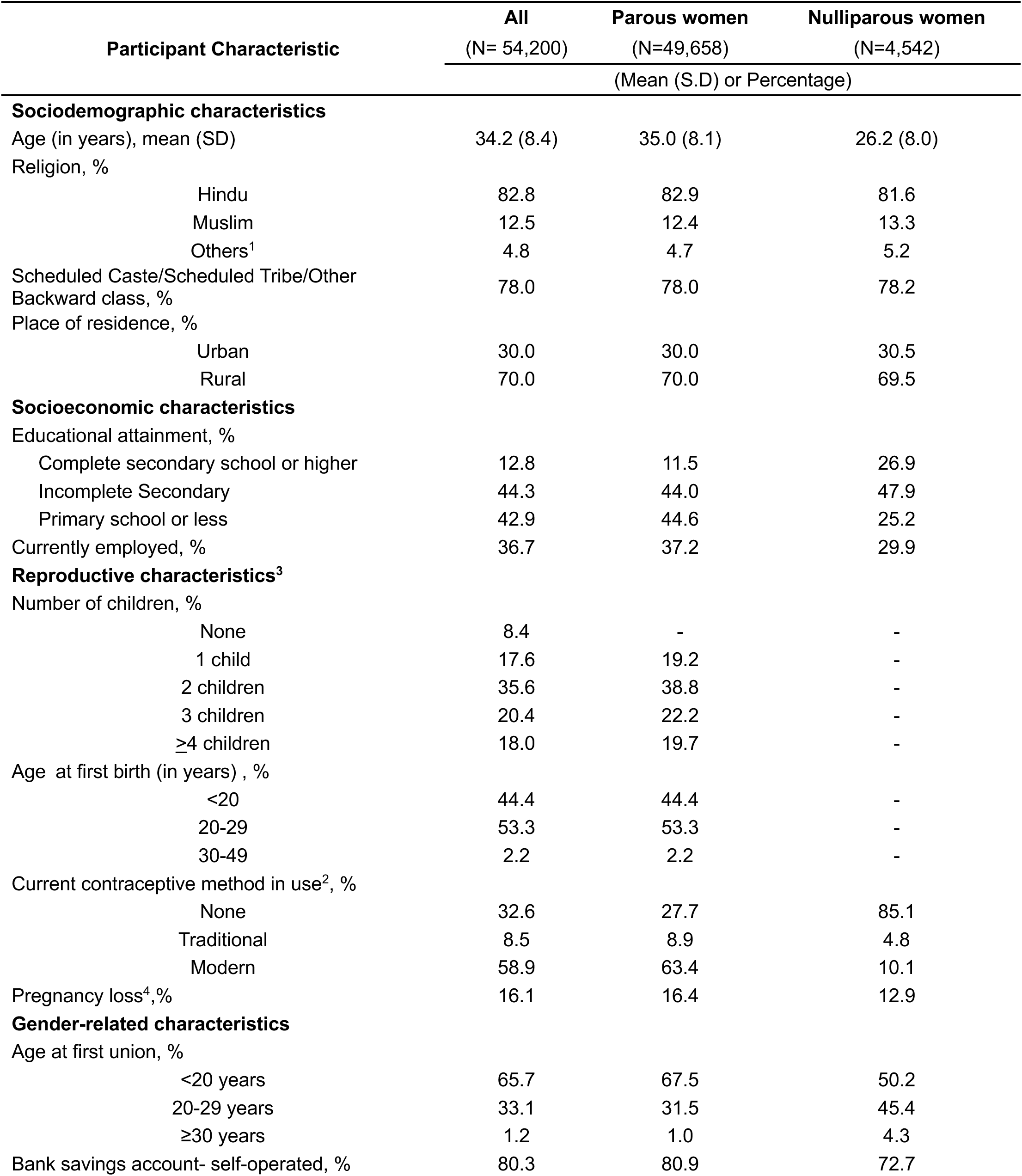
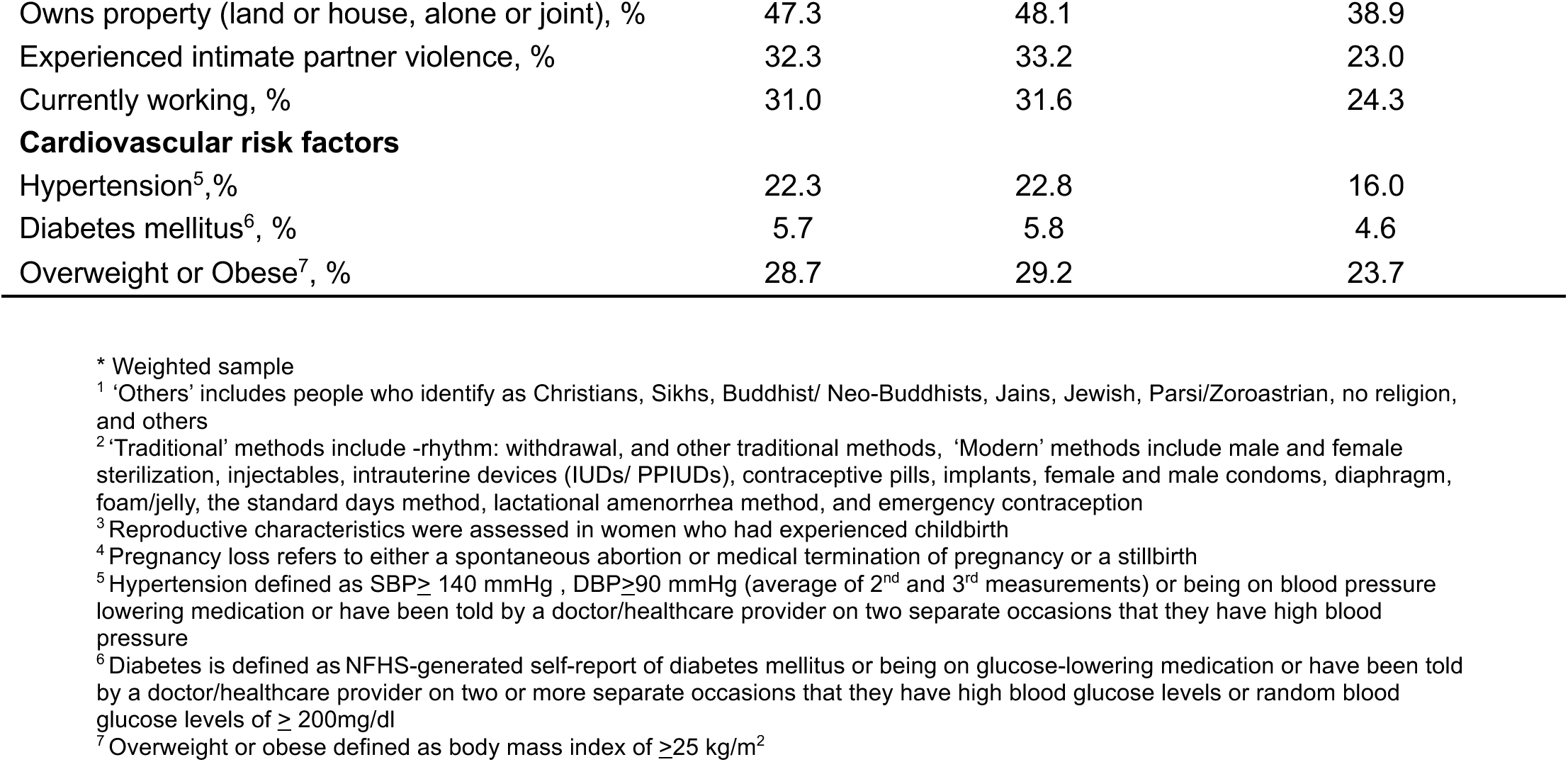
Characteristics of women (18-49 years) in the National Family Health Survey-5*.

### Association between reproductive characteristics and cardiovascular risk factors

Associations between reproductive characteristics and CVD risk factors among women who have ever given birth are shown in **Table 2**. A higher prevalence of hypertension was observed among women with age at first birth younger than 20 years (ref: 20-29 years) [PR (95% CI): 1.11 (1.04, 1.19)] after adjusting for sociodemographics. As with hypertension, a higher prevalence of diabetes was seen among women with age at first birth younger than 20 years [PR (95% CI): 1.17 (1.00, 1.37)] in fully adjusted models. In contrast, a lower prevalence of diabetes was observed with the use of modern methods of contraception (ref: no contraceptive use) [PR (95% CI): 0.81 (0.69, 0.95)] after adjusting for sociodemographics. In fully adjusted models, a higher prevalence of being overweight or obese was observed among women with two children (ref: one child) [PR (95%CI): 1.15 (1.07, 1.24)], among women with age at first birth younger than 20 years [PR (95% CI): 1.06 (1.00, 1.12)], and among those who experienced a pregnancy loss (ref: no pregnancy loss) [PR (95% CI): 1.16 (1.09, 1.23)].

**Table 2:** Cross-sectional associations between reproductive characteristics and prevalence of cardiovascular risk factors among parous women (18-49 years), in the NFHS-5 (N=49,658)

| Reproductive characteristics | Hypertension<sup>1</sup> |  |  |  |  |
| --- | --- | --- | --- | --- | --- |
|  | Prevalence (%) | Age-Adjusted PR* |  | Fully-Adjusted PR** |  |
| Number of children |  |  |  |  |  |
| 1 child | 19.6 | Ref | Ref | Ref | Ref |
| 2 children | 22.6 | 0.97 | [0.88, 1.06] | 0.99 | [0.90, 1.10] |
| 3 children | 24.9 | 0.98 | [0.88, 1.08] | 1.00 | [0.90, 1.11] |
| ≥4 children | 27.6 | 0.99 | [0.89, 1.10] | 1.05 | [0.93, 1.18] |
| Age at first birth (years) |  |  |  |  |  |
| <20 | 24.4 | **1.13** | **[1.06, 1.20]** | **1.11** | **[1.04, 1.19]** |
| 20-29 | 22.5 | Ref | Ref | Ref | Ref |
| 30-49 | 26.0 | 0.97 | [0.81, 1.15] | 0.93 | [0.78, 1.12] |
| Current method of contraception |  |  |  |  |  |
| None | 22.9 | Ref | Ref | Ref | Ref |
| Traditional | 23.3 | 1.03 | [0.92, 1.15] | 1.04 | [0.93, 1.17] |
| Modern | 23.8 | 0.94 | [0.88, 1.01] | 0.97 | [0.90, 1.04] |
| Pregnancy loss |  |  |  |  |  |
| No | 23.7 | Ref | Ref | Ref | Ref |
| Yes | 22.2 | 0.94 | [0.86, 1.03] | 0.92 | [0.85, 1.01] |
| Reproductive characteristics | Diabetes mellitus<sup>2</sup> |  |  |  |  |
|  | Prevalence (%) | Age-Adjusted PR* |  | Fully-Adjusted PR** |  |
| Number of children |  |  |  |  |  |
| 1 child | 4.9 | Ref | Ref | Ref | Ref |
| 2 children | 5.8 | 0.87 | [0.71, 1.07] | 0.92 | [0.74, 1.14] |
| 3 children | 6.3 | 0.8 | [0.64, 1.00] | 0.96 | [0.75, 1.22] |
| ≥4 children | 7.0 | **0.77** | **[0.61, 0.96]** | 1.04 | [0.81, 1.34] |
| Age at first birth (years) |  |  |  |  |  |
| <20 | 6 | 1.09 | [0.94, 1.25] | **1.17** | **[1.00, 1.37]** |
| 20-29 | 5.9 | Ref | Ref | Ref | Ref |
| 30-49 | 7.2 | 0.92 | [0.68, 1.24] | 0.89 | [0.64, 1.23] |
| Current method of contraception |  |  |  |  |  |
| None | 6.1 | Ref | Ref | Ref | Ref |
| Traditional | 7 | 1.21 | [0.91, 1.60] | 1.15 | [0.84, 1.56] |
| Modern | 5.8 | **0.82** | **[0.71, 0.95]** | **0.81** | **[0.69, 0.95]** |
| Pregnancy loss |  |  |  |  |  |
| No | 5.9 | Ref | Ref | Ref | Ref |
| Yes | 6.3 | 1.08 | [0.90, 1.29] | 1.01 | [0.84, 1.21] |

| Reproductive characteristics | Overweight or Obese<sup>3</sup> |  |  |  |  |
| --- | --- | --- | --- | --- | --- |
|  | Prevalence (%) | Age-Adjusted PR* |  | Fully-Adjusted PR** |  |
| Number of children |  |  |  |  |  |
| 1 child | 25.2 | Ref | Ref | Ref | Ref |
| 2 children | 33.2 | **1.10** | **[1.03, 1.19]** | **1.15** | **[1.07, 1.24]** |
| 3 children | 28.6 | **0.87** | **[0.80, 0.95]** | 1.08 | [0.99, 1.17] |
| ≥4 children | 24.4 | **0.68** | **[0.61, 0.75]** | 1.08 | [0.97, 1.19] |
| Age at first birth (years) |  |  |  |  |  |
| <20 | 27 | 0.92 | [0.87, 0.98] | **1.06** | **[1.00, 1.12]** |
| 20-29 | 30.2 | Ref | Ref | Ref | Ref |
| 30-49 | 35.6 | 1.01 | [0.88, 1.17] | 0.87 | [0.75, 1.02] |
| Current method of contraception |  |  |  |  |  |
| None | 25.8 | Ref | Ref | Ref | Ref |
| Traditional | 28.7 | **1.13** | **[1.03, 1.23]** | 1.08 | [0.99, 1.18] |
| Modern | 30.3 | **1.08** | **[1.02, 1.15]** | 1.02 | [0.96, 1.08] |
| Pregnancy loss |  |  |  |  |  |
| No | 28.1 | Ref | Ref | Ref | Ref |
| Yes | 33.2 | **1.19** | **[1.11, 1.26]** | **1.16** | **[1.09, 1.23]** |

Results are presented as Prevalence ratios [95% confidence intervals], Bolded results are statistically significant at p-value<0.05
<sup>1</sup> Hypertension defined as SBP≥140 mmHg, DBP≥90 mmHg (average of 2<sup>nd</sup> and 3<sup>rd</sup> measurements) or on blood pressure lowering medication or have been told by a doctor/healthcare provider on two separate occasions that they have high blood pressure
<sup>2</sup> Diabetes defined as NFHS-generated self-report of diabetes mellitus or on glucose lowering medication or have been told by a doctor/healthcare provider on two or more separate occasions that they have high blood glucose levels
<sup>3</sup> Overweight and Obese defined by a body mass index ≥25 kg/m<sup>2</sup>
\* Age-adjusted model adjusts for age
\*\* Fully-Adjusted model adjusts for age, religion, caste, state, place of residence, wealth index, educational attainment, employment status

### Association between gender-related characteristics and cardiovascular risk factors

Associations between gender-related characteristics and CVD risk factors among parous women are shown in **Table 3**. A greater prevalence of hypertension was observed among women with age at first union younger than 20 years (ref: 20-29 years) [PR (95% CI): 1.09 (1.02, 1.17)] in fully adjusted models. A lower prevalence of diabetes was associated with being currently unemployed (ref: employed) [PR (95% CI): 0.76 (0.58, 1.00)] whereas owning a bank account was associated with a higher prevalence of diabetes [PR (95% CI): 1.22 (1.01, 1.46)]. There were no significant associations between gender-related characteristics and being overweight/obese.

**Table 3:** Cross-sectional associations between gender-related characteristics and prevalence of cardiovascular risk factors among women (18-49 years) in the NFHS-5 (N= 54,200)

| Gender-related characteristics | Hypertension<sup>1</sup> |  |  |  |  |
| --- | --- | --- | --- | --- | --- |
|  | Prevalence (%) | Age-Adjusted PR<sup>*</sup> |  | Fully-Adjusted PR<sup>**</sup> |  |
| Age at first union (years) |  |  |  |  |  |
| <20 | 23.20 | **1.08** | **[1.01, 1.15]** | **1.09** | **[1.02, 1.17]** |
| 20-29 | 21.90 | Ref | Ref | Ref | Ref |
| 30-49 | 33.50 | 1.23 | [0.96, 1.58] | 1.20 | [0.91, 1.58] |
| Have a bank account |  |  |  |  |  |
| Yes | 22.90 | Ref | Ref | Ref | Ref |
| No | 22.70 | 1.05 | [0.97, 1.14] | 1.08 | [0.99, 1.18] |
| Own property (land or home) |  |  |  |  |  |
| Yes | 24.10 | Ref | Ref | Ref | Ref |
| No | 21.80 | 0.97 | [0.92, 1.04] | 0.97 | [0.91, 1.04] |
| Any experience of IPV |  |  |  |  |  |
| Yes | 22.50 | 1.03 | [0.97, 1.10] | 1.03 | [0.97, 1.10] |
| No | 23.80 | Ref | Ref | Ref | Ref |
| Currently working |  |  |  |  |  |
| Yes | 22.50 | Ref | Ref | Ref | Ref |
| No | 23.00 | **1.14** | **[1.07, 1.22]** | 0.99 | [0.88, 1.12] |
| Gender-related characteristics | Diabetes mellitus<sup>2</sup> |  |  |  |  |
|  | Prevalence (%) | Age-Adjusted PR<sup>*</sup> |  | Fully-Adjusted PR<sup>**</sup> |  |
| Age at first union (years) |  |  |  |  |  |
| <20 | 5.80 | 1.02 | [0.89, 1.18] | 1.10 | [0.94, 1.29] |
| 20-29 | 5.80 | Ref | Ref | Ref | Ref |
| 30-49 | 8.70 | 1.06 | [0.59, 1.91] | 1.03 | [0.57, 1.84] |
| Have a bank account |  |  |  |  |  |
| Yes | 5.80 | Ref | Ref | Ref | Ref |
| No | 5.90 | 1.11 | [0.93, 1.33] | **1.22** | **[1.01, 1.46]** |
| Own property (land or home) |  |  |  |  |  |
| Yes | 6.20 | Ref | Ref | Ref | Ref |
| No | 5.50 | 1.00 | [0.87, 1.14] | 0.97 | [0.84, 1.12] |
| Any experience of IPV |  |  |  |  |  |
| Yes | 5.90 | 0.95 | [0.82, 1.10] | 1.02 | [0.87, 1.20] |
| No | 5.80 | Ref | Ref | Ref | Ref |
| Currently working |  |  |  |  |  |
| Yes | 6.10 | Ref | Ref | Ref | Ref |

| No | 5.70 | 1.10 | [0.95, 1.27] | 0.76 | [0.58, 1.00] |
| --- | --- | --- | --- | --- | --- |
| Gender-related characteristics | Overweight or Obese<sup>3</sup> |  |  |  |  |
|  | Prevalence (%) | Age-Adjusted PR* |  | Fully-Adjusted PR** |  |
| Age at first union (years) |  |  |  |  |  |
| <20 | 26.50 | 0.85 | [0.80, 0.89] | 1.03 | [0.98, 1.09] |
| 20-29 | 31.90 | Ref | Ref | Ref | Ref |
| 30-49 | 41.10 | 1.07 | [0.88, 1.30] | 1.01 | [0.80, 1.26] |
| Have a bank account |  |  |  |  |  |
| Yes | 28.90 | Ref | Ref | Ref | Ref |
| No | 26.40 | 0.96 | [0.90, 1.03] | 1.04 | [0.98, 1.11] |
| Own property (land or home) |  |  |  |  |  |
| Yes | 28.70 | Ref | Ref | Ref | Ref |
| No | 28.20 | 1.05 | [1.00, 1.11] | 1.00 | [0.96, 1.06] |
| Any experience of IPV |  |  |  |  |  |
| Yes | 29.70 | 0.85 | [0.81, 0.90] | 0.98 | [0.93, 1.03] |
| No | 25.80 | Ref | Ref | Ref | Ref |
| Currently working |  |  |  |  |  |
| Yes | 27.40 | Ref | Ref | Ref | Ref |
| No | 28.90 | 1.16 | [1.10, 1.23] | 1.08 | [0.98, 1.20] |

Results are presented as Prevalence ratios [95% confidence intervals], Bolded results are statistically significant at p-value<0.05
<sup>1</sup> Hypertension defined as SBP≥ 140 mmHg , DBP≥90 mmHg (average of 2<sup>nd</sup> and 3<sup>rd</sup> measurements) or on blood pressure lowering medication or have been told by a doctor/healthcare provider on two separate occasions that they have high blood pressure
<sup>2</sup> Diabetes defined as NFHS-generated self-report of diabetes mellitus or on glucose lowering medication or have been told by a doctor/healthcare provider on two or more separate occasions that they have high blood glucose levels
<sup>3</sup> Overweight and Obese defined by a body mass index ≥25 kg/m<sup>2</sup>
\* Age-Adjusted model adjusts for age
\*\* Fully-Adjusted model adjusts for age, religion, caste, state, place of residence, wealth index, educational attainment, employment status

### Associations by educational attainment

In the education-stratified analysis for reproductive characteristics, there were no meaningful interactions in parous women (**Table 4**). A younger age at first birth was generally associated with all evaluated CVRF. However, these associations were not always statistically significant. The association between other reproductive characteristics and CVRF across subgroups of educational attainment was similar to the primary analysis. Similarly, the association between gender-related characteristics and CVRF was similar to the primary analysis (**Table 5**). Among women with the lowest educational attainment, there was a statistically significant (p-value for interaction=0.01) greater prevalence of hypertension as compared to other educational attainment groups [PR (95% CI): 1.15 (1.05, 1.25)]. Conversely, there was a statistically significant (p-value for interaction = 0.05) association among women with the highest level of educational attainment, associated with a higher prevalence of diabetes as compared to other subgroups [PR (95% CI): 1.88 (1.16, 3.06)].

**Table 4:** Associations between reproductive characteristics and prevalent cardiovascular risk factors stratified by educational attainment among parous women (18-49 years) in the NFHS-5

| Reproductive characteristics | Hypertension<sup>1</sup> |  |  |  |  |  |  |  |  | p-value for interaction* |
| --- | --- | --- | --- | --- | --- | --- | --- | --- | --- | --- |
|  | Completed secondary school or higher (N=5,697) |  |  | Incomplete secondary school (N=21,836) |  |  | Primary school or less (N= 22,215) |  |  |  |
|  | Prevalence (%) | PR (95% CI) |  | Prevalence (%) | PR (95% CI) |  | Prevalence (%) | PR (95% CI) |  |  |
| Total number of births |  |  |  |  |  |  |  |  |  | 0.69 |
| 1 child | 17.1 | Ref | Ref | 19.9 | Ref | Ref | 21.9 | Ref | Ref |  |
| 2 children | 23.4 | 1.03 | [0.82, 1.30] | 21.4 | 0.95 | [0.83, 1.09] | 24.1 | 0.99 | [0.85, 1.16] |  |
| 3 children | 26.5 | 0.95 | [0.69, 1.29] | 24.5 | 1.0 | [0.86, 1.17] | 25.1 | 0.97 | [0.83, 1.15] |  |
| ≥4 children | 28.2 | 0.99 | [0.62, 1.57] | 28.5 | 1.01 | [0.82, 1.23] | 27.3 | 1.07 | [0.91, 1.25] |  |
| Age at first birth (years) |  |  |  |  |  |  |  |  |  | 0.21 |
| <20 | 19.9 | 1.07 | [0.73, 1.56] | 22.9 | **1.14** | **[1.02, 1.27]** | 25.9 | 1.07 | [0.99, 1.17] |  |
| 20-29 | 20.6 | Ref | Ref | 21.7 | Ref | Ref | 24.6 | Ref | Ref |  |
| 30-49 | 27.5 | 1.02 | [0.74, 1.41] | 27.4 | 1.01 | [0.79, 1.31] | 22.3 | **0.70** | **[0.53, 0.92]** |  |
| Current method of contraception |  |  |  |  |  |  |  |  |  | 0.07 |
| None | 18.8 | Ref | Ref | 21.3 | Ref | Ref | 25.7 | Ref | Ref |  |
| Traditional | 20.6 | 1.12 | [0.82, 1.53] | 25.2 | 1.16 | [0.97, 1.38] | 21.9 | 0.88 | [0.75, 1.04] |  |
| Modern | 22.2 | 1.02 | [0.79, 1.33] | 22.3 | 0.96 | [0.86, 1.06] | 25.5 | 0.95 | [0.86, 1.05] |  |
| Pregnancy loss |  |  |  |  |  |  |  |  |  | 0.07 |
| No | 20.5 | Ref | Ref | 22.8 | Ref | Ref | 25.5 | Ref | Ref |  |
| Yes | 23.6 | 1.08 | [0.87, 1.35] | 20.1 | **0.82** | **[0.72, 0.95]** | 24.2 | 0.97 | [0.86, 1.11] |  |
| Reproductive characteristics | Diabetes mellitus<sup>2</sup> |  |  |  |  |  |  |  |  |  |

|  | Completed secondary school or higher (N=5,697) |  |  | Incomplete secondary school (N=21,836) |  |  | Primary school or less (N= 22,215) |  |  | p-value for interaction* |
| --- | --- | --- | --- | --- | --- | --- | --- | --- | --- | --- |
|  | Prevalence (%) | PR (95% CI) |  | Prevalence (%) | PR (95% CI) |  | Prevalence (%) | PR (95% CI) |  |  |
| Total number of births |  |  |  |  |  |  |  |  |  | 0.17 |
| 1 child | 4.6 | Ref | Ref | 4.8 | Ref | Ref | 5.4 | Ref | Ref |  |
| 2 children | 6.5 | 1.05 | [0.65, 1.71] | 6.1 | 0.96 | [0.69, 1.34] | 5 | 0.72 | [0.52, 1.00] |  |
| 3 children | 4.7 | 0.63 | [0.35, 1.14] | 8.1 | 1.17 | [0.81, 1.69] | 5 | **0.68** | **[0.48, 0.95]** |  |
| ≥4 children | 9.3 | 1.15 | [0.51, 2.57] | 7.8 | 1.08 | [0.70, 1.68] | 6.7 | 0.87 | [0.61, 1.23] |  |
| Age at first birth (years) |  |  |  |  |  |  |  |  |  | 0.43 |
| <20 | 4.4 | 1.10 | [0.62, 1.97] | 6.1 | 1.13 | [0.90, 1.42] | 6 | 1.21 | [0.96, 1.52] |  |
| 20-29 | 5.4 | Ref | Ref | 6.6 | Ref | Ref | 5.2 | Ref | Ref |  |
| 30-49 | 9.1 | 1.16 | [0.61, 2.19] | 5.7 | 0.58 | [0.33, 1.01] | 6.9 | 1.19 | [0.70, 2.02] |  |
| Current method of contraception |  |  |  |  |  |  |  |  |  | 0.09 |
| None | 4.7 | Ref | Ref | 6.2 | Ref | Ref | 6.4 | Ref | Ref |  |
| Traditional | 11.1 | **2.35** | **[1.25, 4.40]** | 6.8 | 1.09 | [0.71, 1.68] | 5.7 | 0.82 | [0.55, 1.21] |  |
| Modern | 5 | 0.99 | [0.63, 1.54] | 6.4 | 0.81 | [0.62, 1.05] | 5.4 | **0.75** | **[0.61, 0.93]** |  |
| Pregnancy loss |  |  |  |  |  |  |  |  |  | 0.69 |
| No | 5.3 | Ref | Ref | 6.4 | Ref | Ref | 5.6 | Ref | Ref |  |
| Yes | 7 | 1.17 | [0.65, 2.10] | 6.4 | 0.99 | [0.76, 1.29] | 5.9 | 0.96 | [0.72, 1.27] |  |
| Overweight or Obese<sup>3</sup> |  |  |  |  |  |  |  |  |  |  |
| Reproductive characteristics | Completed secondary school or higher (N=5,697) |  |  | Incomplete secondary school (N=21,836) |  |  | Primary school or less (N= 22,215) |  |  | p-value for interaction* |
|  | Prevalence (%) | PR (95% CI) |  | Prevalence (%) | PR (95% CI) |  | Prevalence (%) | PR (95% CI) |  |  |
| Total number of births |  |  |  |  |  |  |  |  |  | 0.37 |
| 1 child | 31.7 | Ref | Ref | 24.7 | Ref | Ref | 19 | Ref | Ref |  |
| 2 children | 44.7 | **1.15** | **[1.00, 1.32]** | 35.1 | **1.13** | **[1.02, 1.26]** | 25.5 | **1.21** | **[1.02, 1.42]** |  |
| 3 children | 36.5 | 0.96 | [0.78, 1.19] | 35 | 1.10 | [0.98, 1.25] | 23.5 | 1.09 | [0.92, 1.30] |  |
| ≥4 children | 26.6 | 0.75 | [0.49, 1.14] | 30.8 | 1.00 | [0.86, 1.17] | 22.6 | 1.15 | [0.96, 1.38] |  |
| Age at first birth (years) |  |  |  |  |  |  |  |  |  | 0.79 |
| <20 | 31.5 | 0.98 | [0.77, 1.24] | 30.5 | 1.05 | [0.97, 1.14] | 24.2 | 1.07 | [0.99, 1.17] |  |
| 20-29 | 38.4 | Ref | Ref | 33.3 | Ref | Ref | 22.2 | Ref | Ref |  |
| 30-49 | 45.7 | 0.95 | [0.76, 1.20] | 35.3 | **0.79** | **[0.62, 1.00]** | 24.1 | 0.90 | [0.68, 1.20] |  |
| Current method of contraception |  |  |  |  |  |  |  |  |  | 0.05 |
| None | 37.8 | Ref | Ref | 27.4 | Ref | Ref | 20.6 | Ref | Ref |  |
| Traditional | 33.8 | 0.90 | [0.71, 1.14] | 34.8 | **1.24** | **[1.09, 1.40]** | 19.5 | 0.93 | [0.80, 1.09] |  |
| Modern | 39 | 0.94 | [0.81, 1.09] | 33.8 | 1.04 | [0.95, 1.14] | 24.9 | 1.03 | [0.93, 1.13] |  |
| Pregnancy loss |  |  |  |  |  |  |  |  |  | 0.54 |
| No | 37.2 | Ref | Ref | 30.9 | Ref | Ref | 23.1 | Ref | Ref |  |
| Yes | 42.4 | 1.10 | [0.97, 1.25] | 38.3 | **1.20** | **[1.10, 1.31]** | 24.9 | **1.11** | **[1.00, 1.23]** |  |

Results are presented as Prevalence ratios [95% confidence intervals], Bolded results are statistically significant at p-value<0.05
<sup>1</sup> Hypertension defined as SBP≥ 140 mmHg , DBP≥90 mmHg (average of 2<sup>nd</sup> and 3<sup>rd</sup> measurements) or on blood pressure lowering medication or have been told by a doctor/healthcare provider on two separate occasions that they have high blood pressure
<sup>2</sup> Diabetes defined as NFHS-generated self-report of diabetes mellitus or on glucose lowering medication or have been told by a doctor/healthcare provider on two or more separate occasions that they have high blood glucose levels
<sup>3</sup> Overweight and Obese defined by a body mass index ≥25 kg/m<sup>2</sup>
Model adjusts for age, religion, caste, state, place of residence, wealth index, employment status
\* Interaction between reproductive characteristic and educational attainment

**Table 5:** Cross-sectional associations between gender-related characteristics and prevalence of cardiovascular risk factors among women (18-49 years) in the NFHS-5, stratified by educational attainment

| Gender-related characteristics | Hypertension<sup>1</sup> |  |  |  |  |  |  |  |  | p-value for interaction* |
| --- | --- | --- | --- | --- | --- | --- | --- | --- | --- | --- |
|  | Completed secondary school or higher (N= 6,918) |  |  | Incomplete secondary school (N=24,011) |  |  | Primary school or less (N= 23,271) |  |  |  |
|  | Prevalence (%) | PR (95% CI) |  | Prevalence (%) | PR (95% CI) |  | Prevalence (%) | PR (95% CI) |  |  |
| Age at first union (years) |  |  |  |  |  |  |  |  |  | 0.44 |
| <20 | 18.2 | 1.00 | [0.78, 1.28] | 21.7 | **1.13** | **[1.02, 1.26]** | 24.9 | 1.08 | [0.98, 1.19] |  |
| 20-29 | 20.9 | Ref | Ref | 21.1 | Ref | Ref | 24.5 | Ref | Ref |  |
| 30-49 | 25.5 | 0.90 | [0.60, 1.35] | 33.6 | 1.23 | [0.96, 1.57] | 42.1 | 1.49 | [0.83, 2.66] |  |
| Have a bank account |  |  |  |  |  |  |  |  |  | 0.73 |
| Yes | 20.6 | Ref | Ref | 21.7 | Ref | Ref | 25.1 | Ref | Ref |  |
| No | 18.8 | 1.01 | [0.71, 1.44] | 21.3 | 1.13 | [0.99, 1.29] | 24.5 | 1.06 | [0.95, 1.18] |  |
| Own property (land or home) |  |  |  |  |  |  |  |  |  | 0.05 |
| Yes | 20.9 | Ref | Ref | 22.2 | Ref | Ref | 26.7 | Ref | Ref |  |
| No | 20.0 | 1.09 | [0.89, 1.34] | 21.2 | 1.04 | [0.95, 1.14] | 23.2 | **0.90** | **[0.82, 0.98]** |  |
| Any experience of IPV |  |  |  |  |  |  |  |  |  | **0.01** |
| Yes | 18.6 | 0.89 | [0.70, 1.12] | 20.6 | 0.93 | [0.83, 1.03] | 26.9 | **1.15** | **[1.05, 1.25]** |  |
| No | 20.7 | Ref | Ref | 22.0 | Ref | Ref | 23.7 | Ref | Ref |  |
| Currently working |  |  |  |  |  |  |  |  |  | 0.26 |
| Yes | 23.1 | Ref | Ref | 21.4 | Ref | Ref | 23.3 | Ref | Ref |  |
| No | 19.4 | 1.13 | [0.73, 1.74] | 21.7 | 0.94 | [0.76, 1.17] | 25.9 | 1.03 | [0.88, 1.20] |  |
| Gender-related characteristics | Diabetes mellitus<sup>2</sup> |  |  |  |  |  |  |  |  |  |

|  | Completed secondary school<br>or higher (N= 6,918) |  |  | Incomplete secondary school<br>(N=24,011) |  |  | Primary school or less<br>(N= 23,271) |  |  | p-value for<br>interaction* |
| --- | --- | --- | --- | --- | --- | --- | --- | --- | --- | --- |
|  | Prevalence<br>(%) | PR (95% CI) |  | Prevalence<br>(%) | PR (95% CI) |  | Prevalence<br>(%) | PR (95% CI) |  |  |
| Age at first union (years) |  |  |  |  |  |  |  |  |  | 0.09 |
| <20 | 4.0 | 0.83 | [0.51, 1.36] | 6.0 | 1.17 | [0.94, 1.45] | 5.7 | 1.07 | [0.82, 1.39] |  |
| 20-29 | 5.6 | Ref | Ref | 6.2 | Ref | Ref | 5.4 | Ref | Ref |  |
| 30-49 | 7.4 | 0.77 | [0.39, 1.51] | 13.8 | 1.50 | [0.69, 3.22] | 2.6 | **0.33** | **[0.13, 0.79]** |  |
| Have a bank account |  |  |  |  |  |  |  |  |  | 0.35 |
| Yes | 5.5 | Ref | Ref | 6.1 | Ref | Ref | 5.7 | Ref | Ref |  |
| No | 3.7 | 0.83 | [0.47, 1.45] | 6.6 | **1.39** | **[1.03, 1.88]** | 5.6 | 1.15 | [0.91, 1.45] |  |
| Own property (land or home) |  |  |  |  |  |  |  |  |  | 0.8 |
| Yes | 5.7 | Ref | Ref | 6.6 | Ref | Ref | 6.1 | Ref | Ref |  |
| No | 5.0 | 1.01 | [0.69, 1.48] | 5.9 | 1.01 | [0.82, 1.24] | 5.2 | 0.91 | [0.72, 1.13] |  |
| Any experience of IPV |  |  |  |  |  |  |  |  |  | **0.05** |
| Yes | 8.5 | **1.88** | **[1.16, 3.06]** | 5.7 | 0.93 | [0.71, 1.23] | 5.4 | 0.96 | [0.79, 1.16] |  |
| No | 4.6 | Ref | Ref | 6.4 | Ref | Ref | 5.8 | Ref | Ref |  |
| Currently working |  |  |  |  |  |  |  |  |  | 0.07 |
| Yes | 5.8 | Ref | Ref | 7.4 | Ref | Ref | 5.2 | Ref | Ref |  |
| No | 5.1 | 1.26 | [0.65, 2.44] | 5.7 | 0.68 | [0.42, 1.11] | 5.9 | 0.80 | [0.56, 1.15] |  |
| Overweight or Obese<sup>3</sup> |  |  |  |  |  |  |  |  |  |  |
| Gender-related characteristics | Completed secondary school<br>or higher (N= 6,918) |  |  | Incomplete secondary school<br>(N=24,011) |  |  | Primary school or less<br>(N= 23,271) |  |  | p-value for<br>interaction* |
|  | Prevalence<br>(%) | PR (95% CI) |  | Prevalence<br>(%) | PR (95% CI) |  | Prevalence<br>(%) | PR (95% CI) |  |  |
| Age at first union (years) |  |  |  |  |  |  |  |  |  | 0.29 |
| <20 | 30.6 | 0.97 | [0.83, 1.14] | 30 | **1.08** | **[1.00, 1.16]** | 23.1 | 1.00 | [0.91, 1.09] |  |
| 20-29 | 38.5 | Ref | Ref | 32.5 | Ref | Ref | 23.9 | Ref | Ref |  |
| 30-49 | 42.8 | 0.85 | [0.68, 1.07] | 45.7 | 1.07 | [0.84, 1.36] | 32.7 | 1.18 | [0.59, 2.37] |  |
| Have a bank account |  |  |  |  |  |  |  |  |  | 0.43 |
| Yes | 37.4 | Ref | Ref | 31.1 | Ref | Ref | 23.8 | Ref | Ref |  |
| No | 31.5 | 0.95 | [0.76, 1.17] | 30.7 | 1.08 | [0.99, 1.19] | 21.7 | 1.02 | [0.92, 1.13] |  |
| Own property (land or home) |  |  |  |  |  |  |  |  |  | 0.69 |
| Yes | 36.9 | Ref | Ref | 31.6 | Ref | Ref | 24.1 | Ref | Ref |  |
| No | 36.4 | 1.05 | [0.93, 1.18] | 30.6 | 1.02 | [0.95, 1.09] | 22.6 | 0.96 | [0.89, 1.04] |  |
| Any experience of IPV |  |  |  |  |  |  |  |  |  | 0.65 |
| Yes | 36.6 | 1.02 | [0.88, 1.17] | 28.8 | 0.95 | [0.88, 1.03] | 22.0 | 1.01 | [0.93, 1.09] |  |
| No | 36.7 | Ref | Ref | 32.0 | Ref | Ref | 24.1 | Ref | Ref |  |
| Currently working |  |  |  |  |  |  |  |  |  | 0.12 |
| Yes | 40.5 | Ref | Ref | 30.5 | Ref | Ref | 22.2 | Ref | Ref |  |
| No | 35.3 | 1.14 | [0.89, 1.47] | 31.2 | 0.97 | [0.82, 1.14] | 23.9 | **1.19** | **[1.02, 1.38]** |  |

Results are presented as Prevalence ratios [95% confidence intervals]. Bolded results are statistically significant at p-value<0.05
<sup>1</sup> Hypertension defined as SBP≥ 140 mmHg , DBP≥90 mmHg (average of 2<sup>nd</sup> and 3<sup>rd</sup> measurements) or on blood pressure lowering medication or have been told by a doctor/healthcare provider on two separate occasions that they have high blood pressure
<sup>2</sup> Diabetes defined as NFHS-generated self-report of diabetes mellitus or on glucose lowering medication or have been told by a doctor/healthcare provider on two or more separate occasions that they have high blood glucose levels
<sup>3</sup> Overweight and Obese defined by a body mass index ≥25 kg/m<sup>2</sup>
Model adjusts for age, religion, caste, state, place of residence, wealth index, employment status
\* Interaction between gender-related characteristics and educational attainment

## Discussion

In a large nationally representative sample of Indian women, we found that reproductive and gender-related characteristics were associated with prevalent hypertension, diabetes, and obesity. With respect to reproductive history, women who had their first childbirth before the age of 20 years had a higher prevalence of all assessed CVRFs. Experiencing pregnancy loss was consistently associated with a greater prevalence of being overweight or obese, while the use of modern contraceptives was linked to a lower prevalence of diabetes. On examining gender-related characteristics, an earlier age at first union was positively associated with hypertension. Associations were broadly consistent in direction across socioeconomic subgroups. These findings underscore the importance of considering reproductive life events and gender-related experiences in the evaluation of cardiovascular health across the socioeconomic spectrum in India.

Our study revealed that first childbirth during teenage years was associated with a higher prevalence of CVRF. This adds to prior NFHS-4 findings, which also linked adolescent childbirth to a higher prevalence of hypertension.(48) While prior research suggests a U-shaped relationship between age at first birth and CVD risk, i.e., an increased risk at both younger and older ages, this association has been inconsistent across studies.(17, 49, 50, 51) As demonstrated in high-income countries, the link between reproductive factors and CVD has a strong biological basis.(16, 17) Cardiovascular risk due to a younger age at first birth, however, is thought to have a biological, social, and behavioral basis.(17) Physiologically, adolescent pregnancy triggers unique changes in the body due to the ongoing growth of the adolescent mother, leading to higher weight gain and retention (ref: mothers with older age at first birth), which can increase CVD risk.(17) Their exposure to physiological changes of pregnancy also occurs earlier in life and may irreversibly change their trajectory of cardiovascular risk as compared to women who have childbirth later in life. Socially and behaviorally, adolescent pregnancy can lead to adverse life circumstances, including an incomplete education, reduced economic opportunities, social isolation, and even exposure to violence.(17) These factors contribute to cumulative adversity and increase CVD risk by limiting access to resources and social support while also perpetuating stress and unhealthy living conditions. Together, these interconnected factors suggest that giving birth prior to reaching 20 years of age may negatively impact cardiovascular health through multiple compounding pathways.

We observed conflicting associations between parity and CVD risk factors. While we observed a higher prevalence of being overweight or obese in women with two children (ref: one child), there were no associations between higher parity and diabetes or hypertension. Higher parity is expected to increase CVD risk through repeated physiological changes during pregnancy and the associated metabolic changes.(52) Indeed, several studies in the United States and Britain have demonstrated a J-shaped association between parity and CVD risk (Atherosclerosis Risk in Communities and British Regional Heart Study).(53, 54) Findings from India, however, contrast from these studies and have been inconsistent. In an earlier study using NFHS-4 data, each additional childbirth was associated with significantly lower systolic and diastolic blood pressure, sustained up to 10 years post-childbirth.(55) While sociocultural factors may account for some of the differences, we did not find any meaningful differences in subgroups of educational attainment and occupation status. Additionally, some of our findings could be explained by the cardioprotective effects of breastfeeding, which were unaccounted for,(53) or higher parity in earlier birth cohorts that were less exposed to obesogenic environments. The role of parity in CVD risk warrants further investigation in prospective studies.

In our study, contraceptive use was categorized into traditional and modern methods, with the latter including hormonal methods (e.g.: oral contraceptives) and non-hormonal methods (e.g.: barrier methods, sterilization). Modern contraceptive methods were linked to a lower prevalence of diabetes among parous women, though this association was not always statistically significant. Supplemental analyses (data not shown) showed there were no associations between oral contraceptive pill use and CVRFs, which may be due to the relatively low usage of oral contraception (around 5%) among women in India. Oral contraceptive pills (OCP) increase cardiovascular risk through the atherogenic effects of estrogen exerted through blood pressure and lipid level elevations.(56, 57) Earlier formulations with higher estrogen content were linked to elevated blood pressure, even among normotensive women, with an increase in thromboembolic events.(58, 59) However, more recent analyses challenge these views, suggesting that OCP use might be associated with no or lower risk of cardiovascular events, with prolonged use further reducing these risks.(60, 61) The underlying mechanism contributing to the protective associations observed in our paper is unclear as we could not cleanly parse out the hormonal from the non-hormonal effects.

In this study, pregnancy loss captured stillbirths, spontaneous abortions, and medically terminated pregnancies. Stillbirths and spontaneous abortions, via genetic and immunological pathways, lead to endothelial dysfunction and inflammation, thereby increasing the risk for atherosclerosis, diabetes, and hypertension.(62) In medically terminated pregnancies secondary to maternal health conditions, the increased CVD risk is presumed to be due to the underlying medical issue.(63) Further, increased CVD risk occurs with recurrent pregnancy losses, especially with short intervals.(63, 64, 65, 66, 67) In our analysis, we observed that both parous and nulliparous women who experienced a pregnancy loss were more likely to be overweight or obese. However, we were unable to determine the direction of causality and if there were underlying conditions, such as polycystic ovarian disease, that drove this association.

Gender-related characteristics related to CVRFs have not been studied in the same detail as reproductive characteristics. Conceptually, gender-related characteristics may relate to CVRFs through biological, social, and behavioral pathways. We found that a younger age at first union was associated with a higher prevalence of hypertension. A prior analysis from the India Human Development Survey linked early marriage to a range of health issues in women, including greater functional limitations, poorer self-rated health, and a higher prevalence of diabetes, hypertension, and cardiovascular disease in mid-life.(68) Our study aligns with some of these findings, demonstrating an association between younger age at first marriage and the prevalence of hypertension. The greater prevalence of CVRFs seen with an early age at first marriage could, in part, be attributed to alterations in cardiovascular risk profiles related to pregnancy. Beyond these direct health implications, early marriage adversely impacts educational attainment, correlates with lower empowerment and agency, and is associated with a higher prevalence of intimate partner violence. (69, 70, 71)

Intimate partner violence is a significant factor impacting a woman’s well-being, although the specific mechanisms linking violence with physical health are still under investigation.(72) In our study, women who experienced IPV had a lower prevalence of being overweight or obese.

However, this association did not persist when accounting for sociodemographics. Prior research indicates that early-life abuse—be it physical, sexual, or emotional—increases the risk of developing cardiovascular disease in later life.(73, 74) In adults, physical or sexual IPV is linked to increased abdominal obesity, reduced high-density lipoprotein cholesterol, elevated triglycerides, and greater long-term requirement for anti-hypertensive medication.(75) Notably, one study extended these findings to include both victims and perpetrators of IPV, observing an increased cardiovascular risk in both groups.(76) A comprehensive understanding of the mechanistic pathways linking IPV to CVD risk is needed, particularly its impact at various life stages and its long-term sequelae in later life.

Financial stress negatively impacts health. Our study focused on financial inclusion, measured through the ownership and operation of a bank account and property ownership, individually or jointly. Contrary to our hypothesis that financial inclusion would correlate with better cardiovascular health, we did not find any consistent associations between bank account ownership and/or owning property with CVRFs. Financial inclusion influences health through greater healthcare utilization.(77) Along with property rights, it is pivotal in fostering sustainable livelihoods for women, offering economic security, enhancing their bargaining power, and elevating their status in the community.(78) Our analysis could not assess utilization indicators relevant to cardiovascular disease, such as hospital visits, hospitalization rates, prescription drug use, mortality rates, and patient self-management indicators or medication use, a consideration for future research.

The results of this study should be interpreted considering its strengths and limitations. The study addressed a significant gap in existing research by focusing on gender, reproductive health, and cardiovascular risk in a low- to middle-income country setting. This is particularly important as most existing literature centers on high-income countries, and this study offers insights specific to a different socioeconomic and cultural context. We used a large, comprehensive, nationally representative sample of women in India between 18 and 49 years of age. Given the younger age profile of the study’s participants, there may have been insufficient time for CVRFs to develop within the lifecourse. While the cross-sectional design limits the ability to establish causality, it offers a valuable snapshot of associations between variables in a real-world setting. The study offers insights and directions for subsequent research efforts.

Although the study adjusts for a range of sociodemographic variables, there remains the possibility of residual confounding by behavioral and clinical factors that are known to influence cardiovascular risk. The reliance on self-reported data for several variables, including reproductive history and experiences of intimate partner violence, may introduce reporting biases in the absence of medical records to corroborate the same. Experiences of intimate partner violence are known to be underreported, and it is possible that this was the case in the NFHS as well, which could have resulted in an underestimation of its impact on CVRFs. Another limitation of this study was the separation of sex and gender-related characteristics, which does not accurately reflect their interrelated nature, such as parity and the preference for sons. This is an opportunity for future research to explore how sex and gender influence cardiovascular risk in a more integrated manner.

The findings of this study have both clinical and research implications. Given the high prevalence of early marriage and childbearing among girls and women and the associated health ramifications, healthcare professionals need to identify at-risk individuals for screening and follow-up. Shifting fertility preferences and childbearing patterns in India necessitate an in-depth exploration of the relationship between nulliparity and cardiovascular risk. This exploration should distinguish between women who choose not to have children and those unable to do so for medical reasons. Mechanistic studies that link early age at first marriage and childbirth, pregnancy loss, contraceptive use, and IPV to cardiovascular risk are needed. Lifecourse studies are needed to examine the cumulative effects of adverse social exposures on cardiovascular risk. Given the pervasiveness of IPV, the development of integrated screening protocols in healthcare settings to identify women experiencing IPV and implement interventions to alleviate the associated health and safety risks. Incorporating sex- and gender-specific factors into cardiovascular risk assessment tools and using gender-related and intersectional analyses in research will help better understand health inequities and improve risk evaluation for women.

## Data Availability

The dataset (name: IAIR7ADT) used in the current analysis was obtained through a request to the Demographic and Health Surveys.

## List of abbreviations

CVRF: Cardiovascular risk factor
CVD: Cardiovascular disease
NFHS: National Family Health Survey
SBP: Systolic blood pressure
DBP: Diastolic blood pressure
IPV: Intimate partner violence
SD: Standard deviation
PR: Prevalence ratio
CI: Confidence interval

## Declarations

### Ethics approval and consent to participate

This study was reviewed and approved by the Emory Institutional Review Board under 45 CFR 46.110 and/or 21 CFR 56.110.

### Consent for publication

Not applicable

### Authors’ contributions

VS conceptualized the study, performed the analysis, and prepared the manuscript. SAP and AA provided critical guidance throughout all stages of the research process. KMV, SS, RR, and SY contributed valuable feedback and subject matter expertise to enhance the manuscript. All authors reviewed and approved the final manuscript.

## Acknowledgements

The authors gratefully acknowledge the valuable feedback provided by Dr. Jithin Sam Varghese, Assistant Professor at the Rollins School of Public Health.

## References

1. Gao Z, Chen Z, Sun A, Deng X. Gender differences in cardiovascular disease. Medicine in Novel Technology and Devices. 2019;4:100025.

2. Luijken J, van der Schouw YT, Mensink D, Onland-Moret NC. Association between age at menarche and cardiovascular disease: a systematic review on risk and potential mechanisms. Maturitas. 2017;104:96–116.

3. Norris CM, Yip CY, Nerenberg KA, Clavel MA, Pacheco C, Foulds HJ, et al. State of the science in women’s cardiovascular disease: a Canadian perspective on the influence of sex and gender. Journal of the American Heart Association. 2020;9(4):e015634.

4. Barrett-Connor E. Menopause, atherosclerosis, and coronary artery disease. Current opinion in pharmacology. 2013;13(2):186–91.

5. Connelly PJ, Azizi Z, Alipour P, Delles C, Pilote L, Raparelli V. The importance of gender to understand sex differences in cardiovascular disease. Canadian Journal of Cardiology. 2021;37(5):699–710.

6. O’Neil A, Scovelle AJ, Milner AJ, Kavanagh A. Gender/sex as a social determinant of cardiovascular risk. Circulation. 2018;137(8):854–64.

7. El-Serag R, Thurston RC. Matters of the heart and mind: interpersonal violence and cardiovascular disease in women. Am Heart Assoc; 2020. p. e015479.

8. Chandan JS, Thomas T, Bradbury-Jones C, Taylor J, Bandyopadhyay S, Nirantharakumar K. Risk of cardiometabolic disease and all-cause mortality in female survivors of domestic abuse. Journal of the American Heart Association. 2020;9(4):e014580.

9. Ridgeway CL, Correll SJ. Unpacking the gender system: A theoretical perspective on gender beliefs and social relations. Gender & society. 2004;18(4):510–31.

10. Lips HM. Sex and gender: An introduction: Waveland Press; 2020.

11. Greaves L, Ritz SA. Sex, gender and health: Mapping the landscape of research and policy. International Journal of Environmental Research and Public Health. 2022;19(5):2563.

12. Krieger N. Genders, sexes, and health: what are the connections—and why does it matter? International journal of epidemiology. 2003;32(4):652–7.

13. Murphy E. Estrogen signaling and cardiovascular disease. Circulation research. 2011;109(6):687–96.

14. Willemars MM, Nabben M, Verdonschot JA, Hoes MF. Evaluation of the interaction of sex hormones and cardiovascular function and health. Current Heart Failure Reports. 2022;19(4):200–12.

15. Lim GB. Role of sex hormones in cardiovascular diseases. Nature Reviews Cardiology. 2021;18(6):385-.

16. Khan SS, Cameron NA, Lindley KJ. Pregnancy as an early cardiovascular moment: peripartum cardiovascular health. Circulation Research. 2023;132(12):1584–606.

17. Rosendaal NT, Pirkle CM. Age at first birth and risk of later-life cardiovascular disease: a systematic review of the literature, its limitation, and recommendations for future research. BMC Public Health. 2017;17:1–15.

18. Gill SK. Cardiovascular risk factors and disease in women. Medical Clinics. 2015;99(3):535–52.

19. Lidegaard Ø, Løkkegaard E, Jensen A, Skovlund CW, Keiding N. Thrombotic stroke and myocardial infarction with hormonal contraception. The New England journal of medicine. 2012;366:2257–66.

20. Ley SH, Li Y, Tobias DK, Manson JE, Rosner B, Hu FB, et al. Duration of reproductive life span, age at menarche, and age at menopause are associated with risk of cardiovascular disease in women. Journal of the American Heart Association. 2017;6(11):e006713.

21. Kharazmi E, Fallah M, Luoto R. Maternal age at first delivery and risk of cardiovascular disease later in life. International Scholarly Research Notices. 2013;2013.

22. Palmer JR, Rosenberg L, Shapiro S. Reproductive factors and risk of myocardial infarction. American journal of epidemiology. 1992;136(4):408–16.

23. Ogunmoroti O, Osibogun O, Kolade OB, Ying W, Sharma G, Vaidya D, et al. Multiparity is associated with poorer cardiovascular health among women from the Multi-Ethnic Study of Atherosclerosis. American journal of obstetrics and gynecology. 2019;221(6):631. e1-. e16.

24. Vaidya D, Bennett WL, Sibley CT, Polak JF, Herrington DM, Ouyang P. Association of parity with carotid diameter and distensibility: multi-ethnic study of atherosclerosis. Hypertension. 2014;64(2):253–8.

25. Sanghavi M, Kulinski J, Ayers CR, Nelson D, Stewart R, Parikh N, et al. Association between number of live births and markers of subclinical atherosclerosis: The Dallas Heart Study. European journal of preventive cardiology. 2016;23(4):391–9.

26. Heise L, Greene ME, Opper N, Stavropoulou M, Harper C, Nascimento M, et al. Gender inequality and restrictive gender norms: framing the challenges to health. The Lancet. 2019;393(10189):2440–54.

27. Kennedy E, Binder G, Humphries-Waa K, Tidhar T, Cini K, Comrie-Thomson L, et al. Gender inequalities in health and wellbeing across the first two decades of life: an analysis of 40 low-income and middle-income countries in the Asia-Pacific region. The Lancet Global Health. 2020;8(12):e1473–e88.

28. Doyal L. Sex, gender, and health: the need for a new approach. BMJ (Clinical research ed). 2001;323(7320):1061-3.

29. Vlassoff C. Gender differences in determinants and consequences of health and illness. Journal of health, population, and nutrition. 2007;25(1):47.

30. Mollborn S, Lawrence EM, Hummer RA. A gender framework for understanding health lifestyles. Social Science & Medicine. 2020;265:113182.

31. Eisend M. Gender roles. Journal of Advertising. 2019;48(1):72–80.

32. Garrison-Desany HM, Wilson E, Munos M, Sawadogo-Lewis T, Maïga A, Ako O, et al. The role of gender power relations on women’s health outcomes: evidence from a maternal health coverage survey in Simiyu region, Tanzania. BMC Public Health. 2021;21:1–15.

33. Jejeebhoy SJ, Sathar ZA. Women’s autonomy in India and Pakistan: the influence of religion and region. Population and development review. 2001;27(4):687–712.

34. Subramaniapillai S, Galea LA, Einstein G, de Lange A-MG. Sex and gender in health research: Intersectionality matters. Frontiers in Neuroendocrinology. 2023:101104.

35. Ali MK, Bhaskarapillai B, Shivashankar R, Mohan D, Fatmi ZA, Pradeepa R, et al. Socioeconomic status and cardiovascular risk in urban South Asia: The CARRS Study. European journal of preventive cardiology. 2016;23(4):408–19.

36. Rosengren A, Smyth A, Rangarajan S, Ramasundarahettige C, Bangdiwala SI, AlHabib KF, et al. Socioeconomic status and risk of cardiovascular disease in 20 low-income, middle-income, and high-income countries: the Prospective Urban Rural Epidemiologic (PURE) study. The Lancet Global Health. 2019;7(6):e748–e60.

37. Corsi DJ, Subramanian S. Socioeconomic gradients and distribution of diabetes, hypertension, and obesity in India. JAMA network open. 2019;2(4):e190411-e.

38. Lastrapes WD, Rajaram R. Gender, caste and poverty in India: Evidence from the national family health survey. Eurasian Economic Review. 2016;6:153–71.

39. Fins A. National snapshot: Poverty among women & families, 2020. National Women’s Law Center. 2020.

40. Bhattacharjee S, Hnatkovska V, Lahiri A, editors. The evolution of gender gaps in India. India Policy Forum; 2015: National Council of Applied Economic Research.

41. Evans J, Sahgal N, Salazar AM, Starr KJ, Corichi M. How Indians view gender roles in families and society. Pew Research Center Available online: https://www pewresearchorg/religion/2022/03/02/how-indiansview-gender-roles-in-families-and-society/(accessed on 15 January 2022). 2022.

42. Fund IIfPSaIC. National Family Health Survey (NFHS-5), 2019-21: India: Volume I. Mumbai: International Institute for Population Sciences and Inner City Fund; 2021.

43. Shah SN, Munjal Y, Kamath SA, Wander GS, Mehta N, Mukherjee S, et al. Indian guidelines on hypertension-IV (2019). Journal of human hypertension. 2020;34(11):745–58.

44. Straus MA, Hamby SL, Boney-McCoy S, Sugarman DB. The revised conflict tactics scales (CTS2) development and preliminary psychometric data. Journal of family issues. 1996;17(3):283–316.

45. Lynch J, Kaplan G. Socioeconomic position: Social epidemiology. New York: Oxford University Press; 2000.

46. Krieger N, Williams DR, Moss NE. Measuring social class in US public health research: concepts, methodologies, and guidelines. Annual review of public health. 1997;18(1):341–78.

47. Barros AJ, Hirakata VN. Alternatives for logistic regression in cross-sectional studies: an empirical comparison of models that directly estimate the prevalence ratio. BMC medical research methodology. 2003;3(1):1–13.

48. Datta BK, Husain MJ, Kostova D. Hypertension in women: the role of adolescent childbearing. BMC Public Health. 2021;21(1):1–14.

49. Mirowsky J. Age at first birth, health, and mortality. Journal of health and social behavior. 2005;46(1):32–50.

50. Cooke C-LM, Davidge ST. Advanced maternal age and the impact on maternal and offspring cardiovascular health. American Journal of Physiology-Heart and Circulatory Physiology. 2019;317(2):H387–H94.

51. Wolfson C, Gemmill A, Strobino DM. Advanced maternal age and its association with cardiovascular disease in later life. Women’s Health Issues. 2022;32(3):219–25.

52. Li W, Ruan W, Lu Z, Wang D. Parity and risk of maternal cardiovascular disease: a dose–response meta-analysis of cohort studies. European journal of preventive cardiology. 2019;26(6):592–602.

53. Oliver-Williams C, Vladutiu CJ, Loehr LR, Rosamond WD, Stuebe AM. The association between parity and subsequent cardiovascular disease in women: the atherosclerosis risk in communities study. Journal of women’s health. 2019;28(5):721–7.

54. Lawlor DA, Emberson JR, Ebrahim S, Whincup PH, Wannamethee SG, Walker M, et al. Is the association between parity and coronary heart disease due to biological effects of pregnancy or adverse lifestyle risk factors associated with child-rearing? Findings from the British Women’s Heart and Health Study and the British Regional Heart Study. Circulation. 2003;107(9):1260–4.

55. Teufel F, Geldsetzer P, Sudharsanan N, Subramanyam M, Yapa HM, De Neve J-W, et al. The effect of bearing and rearing a child on blood pressure: a nationally representative instrumental variable analysis of 444 611 mothers in India. International journal of epidemiology. 2021;50(5):1671–83.

56. Tepper NK, Krashin JW, Curtis KM, Cox S, Whiteman MK. Update to CDC’s US medical eligibility criteria for contraceptive use, 2016: revised recommendations for the use of hormonal contraception among women at high risk for HIV infection. Morbidity and Mortality Weekly Report. 2017;66(37):990.

57. Lindley KJ, Bairey Merz CN, Davis MB, Madden T, Park K, Bello NA, et al. Contraception and reproductive planning for women with cardiovascular disease: JACC focus seminar 5/5. Journal of the American College of Cardiology. 2021;77(14):1823–34.

58. Shufelt C, LeVee A. Hormonal contraception in women with hypertension. Jama. 2020;324(14):1451–2.

59. Lidegaard Ø, Nielsen LH, Skovlund CW, Skjeldestad FE, Løkkegaard E. Risk of venous thromboembolism from use of oral contraceptives containing different progestogens and oestrogen doses: Danish cohort study, 2001-9. BMJ (Clinical research ed). 2011;343.

60. Dou W, Huang Y, Liu X, Huang C, Huang J, Xu B, et al. Associations of Oral Contraceptive Use With Cardiovascular Disease and All-Cause Death: Evidence From the UK Biobank Cohort Study. Journal of the American Heart Association. 2023;12(16):e030105.

61. Brabaharan S, Veettil SK, Kaiser JE, Rao VRR, Wattanayingcharoenchai R, Maharajan M, et al. Association of hormonal contraceptive use with adverse health outcomes: an umbrella review of meta-analyses of randomized clinical trials and cohort studies. JAMA network open. 2022;5(1):e2143730-e.

62. Ranthe MF, Andersen EAW, Wohlfahrt J, Bundgaard H, Melbye M, Boyd HA. Pregnancy loss and later risk of atherosclerotic disease. Circulation. 2013;127(17):1775–82.

63. Kyriacou H, Al-Mohammad A, Muehlschlegel C, Foster-Davies L, Bruco MEF, Legard C, et al. The risk of cardiovascular diseases after miscarriage, stillbirth, and induced abortion: a systematic review and meta-analysis. European Heart Journal Open. 2022;2(5):oeac065.

64. Smith GC, Pell JP, Walsh D. Spontaneous loss of early pregnancy and risk of ischaemic heart disease in later life: retrospective cohort study. BMJ (Clinical research ed). 2003;326(7386):423-4.

65. Oliver-Williams CT, Heydon EE, Smith GC, Wood AM. Miscarriage and future maternal cardiovascular disease: a systematic review and meta-analysis. Heart. 2013.

66. Peters SA, Yang L, Guo Y, Chen Y, Bian Z, Tian X, et al. Pregnancy, pregnancy loss, and the risk of cardiovascular disease in Chinese women: findings from the China Kadoorie Biobank. BMC medicine. 2017;15:1–10.

67. Okoth K, Subramanian A, Chandan JS, Adderley NJ, Thomas GN, Nirantharakumar K, et al. Long term miscarriage-related hypertension and diabetes mellitus. Evidence from a United Kingdom population-based cohort study. PloS one. 2022;17(1):e0261769.

68. Vikram K. Early marriage and health among women at midlife: Evidence from India. Journal of Marriage and Family. 2021;83(5):1480–501.

69. Marphatia AA, Ambale GS, Reid AM. Women’s marriage age matters for public health: a review of the broader health and social implications in South Asia. Frontiers in public health. 2017;5:269.

70. Roychowdhury P, Dhamija G. The causal impact of women’s age at marriage on domestic violence in india. Fem Econ. 2021;27(3):188–220.

71. Santhya KG, Ram U, Acharya R, Jejeebhoy SJ, Ram F, Singh A. Associations between early marriage and young women’s marital and reproductive health outcomes: evidence from India. International perspectives on sexual and reproductive health. 2010:132–9.

72. Liu X, Logan J, Alhusen J. Cardiovascular risk and outcomes in women who have experienced intimate partner violence: an integrative review. Journal of cardiovascular nursing. 2020;35(4):400–14.

73. Wegman HL, Stetler C. A meta-analytic review of the effects of childhood abuse on medical outcomes in adulthood. Psychosomatic medicine. 2009;71(8):805–12.

74. Suglia SF, Sapra KJ, Koenen KC. Violence and cardiovascular health: a systematic review. Am J Prev Med. 2015;48(2):205–12.

75. Stene LE, Jacobsen GW, Dyb G, Tverdal A, Schei B. Intimate partner violence and cardiovascular risk in women: a population-based cohort study. Journal of women’s health. 2013;22(3):250–8.

76. Clark CJ, Alonso A, Everson-Rose SA, Spencer RA, Brady SS, Resnick MD, et al. Intimate partner violence in late adolescence and young adulthood and subsequent cardiovascular risk in adulthood. Preventive medicine. 2016;87:132–7.

77. Singh A, Kumar K, McDougal L, Silverman JG, Atmavilas Y, Gupta R, et al. Does owning a bank account improve reproductive and maternal health services utilization and behavior in India? Evidence from the National Family Health Survey 2015–16. SSM-Population Health. 2019;7.

78. Steinzor N, editor Women’s Property and Inheritance Rights: Improving Lives in a Changing Time. Final Synthesis and Conference Proceedings Paper Washington: WIDTECH and Development Alternatives, Inc; 2003.

