## Supplement for "Association of reproductive and gender-related characteristics with cardiovascular risk factors of women in India: an analysis of nationally representative data"

### Supplemental Methods

National Family Health Survey-5 sampling strategy

Primary sampling units (PSU) were systematically selected across rural and urban strata. Rural strata were further divided based on the estimated number of households and the proportions of scheduled caste and scheduled tribe populations. PSUs were then organized by literacy rates among women/girls over six years of age, with final PSU selections done through probability proportional to size systematic sampling.[1] In urban areas, a parallel process was implemented. Census enumeration blocks were sorted by scheduled caste/tribe populations, followed by probability proportional to size systematic sampling selection.[1] Subsequent to PSU selection, comprehensive household mapping and listing processes were conducted. Larger PSUs, comprising at least 300 households, were divided into smaller groups of 100-150 households. From these, two segments were randomly chosen. Within each segment, 22 households were systematically sampled. Participants for the NFHS-5 were then selected from these households, ensuring a comprehensive and representative data collection process.

### Statistical analysis

To clarify the analytic sample in models evaluating age at first birth and first union with cardiovascular risk factors, in supplemental analyses, we stratified by current age so that categories were relevant (e.g., among women who were currently under 20 years of age, we only evaluated the association of age at first birth under 20 years with cardiovascular risk factors). In an additional analysis, we restricted to women who were between 30 and 49 years old and evaluated the association between age at first birth/ union with cardiovascular risk factors.

### Supplemental Results

Association of reproductive characteristics with cardiovascular risk factors in nulliparous women

Among nulliparous women (**Supplemental Table 1**), experiencing a pregnancy loss was associated with a higher prevalence of being overweight or obese [PR (95% CI): 1.46 (1.17, 1.82)] in fully adjusted models. There were no significant interactions between educational attainment and reproductive characteristics in nulliparous women (**Supplemental Table 2**).

Association of reproductive characteristics with cardiovascular risk factors in parous women- Stratification and Restriction by current age

On restricting our sample to women aged 30-49 years (**Supplemental Table 3**), an age at first birth younger than 20 years (ref: 20-29 years) was associated with a greater prevalence of all evaluated risk factors in the fully adjusted models. We did not see consistent associations between age at first union and any of the cardiovascular risk factors.

On stratifying our sample by current age (**Supplemental Table 4**), we found that women who were currently younger than 20 years and had experienced childbirth or union had a lower prevalence of being overweight or obese in fully adjusted models. However, women aged 30 years or older had a higher prevalence of being overweight or obese irrespective of age at first birth.

**Supplemental Table 1: Cross-sectional associations between reproductive characteristics and prevalence of cardiovascular risk factors among women who have not experienced childbirth (18-49 years) in the NFHS-5 (N=4,542)**

| Reproductive characteristics | Hypertension[1] | | | | |
|---|---|---|---|---|---|
| | Prevalence (%) | Age-Adjusted PR[*] | | Fully-Adjusted PR[**] | |
| Current method of contraception | | | | | |
| None | 17.1 | Ref | Ref | Ref | Ref |
| Traditional | 11.5 | 0.72 | [0.44, 1.17] | 0.67 | [0.41, 1.09] |
| Modern | 14.2 | 0.95 | [0.63, 1.41] | 0.92 | [0.63, 1.34] |
| Ever terminated a pregnancy | | | | | |
| No | 15.8 | Ref | Ref | Ref | Ref |
| Yes | 21.3 | 1.21 | [0.87, 1.67] | 1.20 | [0.86, 1.67] |
| **Reproductive characteristics** | **Diabetes mellitus[2]** | | | | |
| | **Prevalence (%)** | **Age-Adjusted PR[*]** | | **Fully-Adjusted PR[**]** | |
| Current method of contraception | | | | | |
| None | 4.8 | Ref | Ref | Ref | Ref |
| Traditional | 2 | 0.57 | [0.18, 1.84] | 0.51 | [0.15, 1.71] |
| Modern | 3.4 | 0.81 | [0.37, 1.74] | 0.80 | [0.37, 1.75] |
| Pregnancy loss | | | | | |
| No | 4.3 | Ref | Ref | Ref | Ref |
| Yes | 6.1 | 1.19 | [0.70, 2.04] | 1.17 | [0.69, 1.99] |
| **Reproductive characteristics** | **Overweight or Obese[3]** | | | | |
| | **Prevalence (%)** | **Age-Adjusted PR[*]** | | **Fully-Adjusted PR[**]** | |
| Current method of contraception | | | | | |
| None | 24.2 | Ref | Ref | Ref | Ref |
| Traditional | 16 | 0.81 | [0.51, 1.29] | 0.72 | [0.45, 1.16] |
| Modern | 19.6 | 0.89 | [0.61, 1.30] | 0.78 | [0.53, 1.15] |
| Pregnancy loss | | | | | |
| No | 21.8 | Ref | Ref | Ref | Ref |
| Yes | 34.2 | **1.41** | **[1.12, 1.78]** | **1.46** | **[1.17, 1.82]** |

Results are presented as Prevalence ratios [95% confidence intervals], Bolded results are statistically significant at p-value<0.05
[1] Hypertension defined as SBP$\geq$ 140 mmHg , DBP$\geq$90 mmHg (average of 2nd and 3rd measurements) or on blood pressure lowering medication or have been told by a doctor/healthcare provider on two separate occasions that they have high blood pressure
[2] Diabetes defined as NFHS-generated self-report of diabetes mellitus or on glucose lowering medication or have been told by a doctor/healthcare provider on two or more separate occasions that they have high blood glucose levels
[3] Overweight and Obese defined by a body mass index $\geq$25 kg/$m^2$
[*] Age-adjusted model adjusts for age
[**] Fully-adjusted model adjusts for age, religion, caste, state, place of residence, wealth index, educational attainment, employment status

117

**Supplemental Table 2: Cross-sectional associations between reproductive characteristics and prevalence of cardiovascular risk factors among nulliparous women (18-49 years) in the NFHS-5, stratified by educational attainment**

| Reproductive characteristics | Hypertension[1] | | | | | | | | | p-value for interaction* |
|---|---|---|---|---|---|---|---|---|---|---|
| | Completed secondary school or higher (N= 1,221) | | | Incomplete secondary school (N=2,175) | | | Primary school or less (N=1,146) | | | |
| | Prevalence (%) | PR (95% CI) | | Prevalence (%) | PR (95% CI) | | Prevalence (%) | PR (95% CI) | | |
| Current method of contraception | | | | | | | | | | 0.54 |
| None | 18.0 | Ref | Ref | 15.2 | Ref | Ref | 19.7 | Ref | Ref | |
| Traditional | 11.9 | 0.39 | [0.13, 1.19] | 13.0 | 0.97 | [0.54, 1.74] | 6.8 | 0.56 | [0.21, 1.51] | |
| Modern | 16.3 | 1.14 | [0.57, 2.28] | 12.1 | 0.83 | [0.51, 1.35] | 14.3 | 0.85 | [0.39, 1.85] | |
| Pregnancy loss | | | | | | | | | | 0.14 |
| No | 15.2 | Ref | Ref | 14.6 | Ref | Ref | 18.9 | Ref | Ref | |
| Yes | 33.8 | **1.90** | **[1.08, 3.34]** | 16.4 | 0.99 | [0.64, 1.54] | 18.3 | 0.87 | [0.49, 1.55] | |

| Reproductive characteristics | Diabetes mellitus[2] | | | | | | | | | p-value for interaction* |
|---|---|---|---|---|---|---|---|---|---|---|
| | Completed secondary school or higher (N= 1,221) | | | Incomplete secondary school (N=2,175) | | | Primary school or less (N=1,146) | | | |
| | Prevalence (%) | PR (95% CI) | | Prevalence (%) | PR (95% CI) | | Prevalence (%) | PR (95% CI) | | |
| Current method of contraception | | | | | | | | | | 0.11 |
| None | 4.5 | Ref | Ref | 4.7 | Ref | Ref | 5.3 | Ref | Ref | |
| Traditional | 1.1 | 0.21 | [0.03, 1.69] | 1.5 | 0.47 | [0.07, 3.19] | 4.7 | 1.44 | [0.23, 9.00] | |
| Modern | 1.6 | 0.37 | [0.07, 1.97] | 1.7 | 0.44 | [0.15, 1.27] | 11.4 | 2.37 | [0.78, 7.16] | |
| Pregnancy loss | | | | | | | | | | 0.63 |
| No | 3.4 | Ref | Ref | 4.3 | Ref | Ref | 5.4 | Ref | Ref | |
| Yes | 7.2 | 1.62 | [0.47, 5.63] | 4.4 | 0.81 | [0.30, 2.15] | 7.8 | 1.15 | [0.42, 3.18] | |

| Reproductive characteristics | Overweight or Obese[3] | | | | | | | | | p-value for interaction* |
|---|---|---|---|---|---|---|---|---|---|---|
| | Completed secondary school or higher (N= 1,221) | | | Incomplete secondary school (N=2,175) | | | Primary school or less (N=1,146) | | | |
| | Prevalence (%) | PR (95% CI) | | Prevalence (%) | PR (95% CI) | | Prevalence (%) | PR (95% CI) | | |
| Current method of contraception | | | | | | | | | | 0.09 |
| None | 32.0 | Ref | Ref | 21.5 | Ref | Ref | 22.1 | Ref | Ref | |
| Traditional | 22.2 | 0.80 | [0.39, 1.64] | 12.7 | 0.59 | [0.29, 1.17] | 14.6 | 0.94 | [0.36, 2.42] | |
| Modern | 22.5 | 0.71 | [0.40, 1.26] | 9.9 | **0.48** | **[0.30, 0.78]** | 35.6 | 1.70 | [0.78, 3.72] | |
| Pregnancy loss | | | | | | | | | | 0.54 |
| No | 28.9 | Ref | Ref | 18.6 | Ref | Ref | 20.2 | Ref | Ref | |
| Yes | 37.5 | 1.19 | [0.77, 1.84] | 31.1 | **1.47** | **[1.04, 2.08]** | 35.9 | 1.54 | [0.92, 2.59] | |

Results are presented as Prevalence ratios [95% confidence intervals], Bolded results are statistically significant at p-value<0.05
[1] Hypertension defined as SBP≥ 140 mmHg , DBP≥90 mmHg (average of 2nd and 3rd measurements) or on blood pressure lowering medication or have been told by a doctor/healthcare provider on two separate occasions that they have high blood pressure
[2] Diabetes defined as NFHS-generated self-report of diabetes mellitus or on glucose-lowering medication or have been told by a doctor/healthcare provider on two or more separate occasions that they have high blood glucose levels
[3] Overweight and Obese defined by a body mass index ≥25 $kg/m^2$
The model adjusts for age, religion, and caste as the sample size was too small to generate estimates with confidence intervals using the fully-adjusted models used for parous women.
* Interaction between reproductive characteristic and educational attainment

**Supplemental Table 3**: Cross-sectional associations between age at first union and childbirth with the prevalence of cardiovascular risk factors among women (30-49 years), in the NFHS-5

| | Hypertension[1] | | | | |
|---|---|---|---|---|---|
| | Prevalence (%) | Age-Adjusted PR* | | Fully-Adjusted PR** | |
| Age at first birth (years)* (N=37,234) | | | | | |
| <20 | 27.2 | **1.09** | **[1.01, 1.17]** | **1.12** | **[1.04, 1.20]** |
| 20-29 | 25.1 | Ref | Ref | Ref | Ref |
| 30-49 | 26.0 | 1.04 | [0.86, 1.24] | 0.91 | [0.76, 1.10] |
| Age at first union (years) (N=38,524) | | | | | |
| <20 | 26.1 | **1.08** | **[1.00, 1.17]** | 1.03 | [0.96, 1.11] |
| 20-29 | 25.4 | Ref | Ref | Ref | Ref |
| 30-49 | 37.8 | 1.18 | [0.90, 1.55] | **1.38** | **[1.04, 1.82]** |
| | **Diabetes mellitus[2]** | | | | |
| | **Prevalence (%)** | **Age-Adjusted PR*** | | **Fully-Adjusted PR**** | |
| Age at first birth (years) (N=37,234) | | | | | |
| <20 | 7.4 | 1.09 | [0.93, 1.27] | **1.22** | **[1.03, 1.45]** |
| 20-29 | 6.8 | Ref | Ref | Ref | Ref |
| 30-49 | 7.5 | 1.09 | [0.80, 1.48] | 0.88 | [0.63, 1.23] |
| Age at first union (years) (N=38,524) | | | | | |
| <20 | 7.0 | 1.11 | [0.94, 1.32] | 0.98 | [0.83, 1.15] |
| 20-29 | 7.3 | Ref | Ref | Ref | Ref |
| 30-49 | 7.3 | 1.01 | [0.57, 1.81] | 1.26 | [0.68, 2.32] |
| | **Overweight or Obese[3]** | | | | |
| | **Prevalence (%)** | **Age-Adjusted PR*** | | **Fully-Adjusted PR**** | |
| Age at first birth (years) (N=37,234) | | | | | |
| <20 | 32.4 | **0.93** | **[0.88, 0.99]** | **1.07** | **[1.01, 1.14]** |
| 20-29 | 34.7 | Ref | Ref | Ref | Ref |
| 30-49 | 34.5 | 0.99 | [0.85, 1.15] | 0.88 | [0.75, 1.02] |
| Age at first union (years) (N=38,524) | | | | | |
| <20 | 31.9 | 1.04 | [0.98, 1.11] | **0.86** | **[0.81, 0.91]** |
| 20-29 | 37.1 | Ref | Ref | Ref | Ref |
| 30-49 | 39.8 | 1.02 | [0.81, 1.29] | 1.10 | [0.88, 1.37] |

* Among parous women

Results are presented as Prevalence ratios [95% confidence intervals], Bolded results are statistically significant at p-value<0.05

[1] Hypertension defined as SBP$\geq$140 mmHg , DBP$\geq$90 mmHg (average of 2nd and 3rd measurements) or on blood pressure lowering medication or have been told by a doctor/healthcare provider on two separate occasions that they have high blood pressure

[2] Diabetes defined as NFHS-generated self-report of diabetes mellitus or on glucose lowering medication or have been told by a doctor/healthcare provider on two or more separate occasions that they have high blood glucose levels

[3] Overweight and Obese defined by a body mass index $\geq$25 $kg/m^2$

* Age-adjusted model adjusts for age

** Fully-Adjusted model adjusts for age, religion, caste, state, place of residence, wealth index, educational attainment, employment status

**Supplemental Table 4**: Cross-sectional associations between age at first union and childbirth with the prevalence of cardiovascular risk factors among women (18-49 years), in the NFHS-5

| | Hypertension[1] | | | | |
|---|---|---|---|---|---|
| | Prevalence (%) | Age-Adjusted PR[*] | | Fully-Adjusted PR[**] | |
| **Age at first birth (years)*** (N=52,877) | | | | | |
| age at 1st birth 20-29, current age 20-29 (n=8751) | 14.2 | Ref | Ref | Ref | Ref |
| age at 1st birth <20, current age <20 (n=367) | 16.1 | 1.13 | [0.71, 1.80] | 1.46 | [0.92, 2.30] |
| age at 1st birth <20, current age 20-29 (n=6525) | 15.3 | 1.07 | [0.92, 1.25] | 1.11 | [0.96, 1.29] |
| age at 1st birth <20, current age ≥30 (n=13983) | 27.2 | **1.91** | **[1.72, 2.13]** | 1.08 | [0.94, 1.24] |
| age at 1st birth 20-29, current age ≥30 (n=21686) | 25.1 | **1.76** | **[1.59, 1.95]** | 0.98 | [0.85, 1.12] |
| age at 1st birth >=30, current age ≥30 (n=1565) | 26.0 | **1.83** | **[1.49, 2.23]** | 0.91 | [0.73, 1.14] |
| **Age at first union (years)** (N=56,662) | | | | | |
| age at 1st union 20-29, current age 20-29 (n=6581) | 13.3 | Ref | Ref | Ref | Ref |
| age at 1st union <20, current age <20 (n=367) | 16.1 | 1.19 | [0.74, 1.92] | 1.56 | [0.98, 2.48] |
| age at 1st union <20, current age 20-29 (n=11190) | 15.3 | 1.12 | [0.97, 1.31] | **1.20** | **[1.03, 1.40]** |
| age at 1st union <20, current age ≥30 (n=23103) | 26.1 | **1.95** | **[1.72, 2.20]** | 1.12 | [0.96, 1.31] |
| age at 1st union 20-29, current age ≥30 (n=14480) | 25.4 | **1.89** | **[1.66, 2.14]** | 1.05 | [0.90, 1.23] |
| age at 1st union >=30, current age ≥30 (n=941) | 37.8 | **2.59** | **[1.94, 3.47]** | 1.25 | [0.93, 1.67] |
| | **Diabetes mellitus[2]** | | | | |
| | **Prevalence (%)** | **Age-Adjusted PR[*]** | | **Fully-Adjusted PR[**]** | |
| **Age at first birth (years)*** (N=52,877) | | | | | |
| age at 1st birth 20-29, current age 20-29 (n=8751) | 2.8 | Ref | Ref | Ref | Ref |
| age at 1st birth <20, current age <20 (n=367) | 0.8 | 0.28 | [0.08, 0.99] | 0.49 | [0.14, 1.76] |
| age at 1st birth <20, current age 20-29 (n=6525) | 2.5 | 0.89 | [0.58, 1.39] | 1.01 | [0.65, 1.58] |
| age at 1st birth <20, current age ≥30 (n=13983) | 7.4 | **2.66** | **[1.94, 3.63]** | 1.00 | [0.67, 1.49] |
| age at 1st birth 20-29, current age ≥30 (n=21686) | 6.8 | **2.44** | **[1.79, 3.33]** | 0.83 | [0.57, 1.22] |
| age at 1st birth >=30, current age ≥30 (n=1565) | 7.5 | **2.66** | **[1.77, 4.00]** | 0.75 | [0.46, 1.22] |
| **Age at first union (years)** (N=56,662) | | | | | |
| age at 1st union 20-29, current age 20-29 (n=6581) | 2.7 | Ref | Ref | Ref | Ref |
| age at 1st union <20, current age <20 (n=367) | 0.8 | **0.28** | **[0.08, 0.99]** | 0.49 | [0.14, 1.77] |
| age at 1st union <20, current age 20-29 (n=11190) | 2.7 | 0.97 | [0.66, 1.44] | 1.14 | [0.76, 1.70] |
| age at 1st union <20, current age ≥30 (n=23103) | 7.0 | **2.51** | **[1.84, 3.42]** | 0.99 | [0.67, 1.48] |
| age at 1st union 20-29, current age ≥30 (n=14480) | 7.3 | **2.57** | **[1.85, 3.58]** | 0.91 | [0.60, 1.37] |
| age at 1st union >=30, current age ≥30 (n=941) | 7.3 | **3.23** | **[1.66, 6.31]** | 0.93 | [0.47, 1.83] |
| | **Overweight or Obese[3]** | | | | |
| | **Prevalence (%)** | **Age-Adjusted PR[*]** | | **Fully-Adjusted PR[**]** | |
| **Age at first birth (years)*** (N=52,877) | | | | | |
| age at 1st birth 20-29, current age 20-29 (n=8751) | 20.3 | Ref | Ref | Ref | Ref |
| age at 1st birth <20, current age <20 (n=367) | 4.1 | **0.20** | **[0.10, 0.43]** | **0.28** | **[0.13, 0.58]** |
| age at 1st birth <20, current age 20-29 (n=6525) | 16.0 | **0.79** | **[0.69, 0.90]** | 0.96 | [0.85, 1.10] |
| age at 1st birth <20, current age ≥30 (n=13983) | 32.4 | **1.60** | **[1.45, 1.76]** | **1.43** | **[1.28, 1.61]** |

| | | | | | |
|---|---|---|---|---|---|
| age at 1st birth 20-29, current age ≥30 (n=21686) | 34.7 | **1.72** | **[1.56, 1.88]** | **1.32** | **[1.19, 1.48]** |
| age at 1st birth >=30, current age ≥30 (n=1565) | 34.5 | **1.70** | **[1.44, 2.01]** | 1.15 | [0.95, 1.38] |
| **Age at first union (years)** (N=56,662) | | | | | |
| age at 1st union 20-29, current age 20-29 (n=6581) | 21.8 | Ref | Ref | Ref | Ref |
| age at 1st union <20, current age <20 (n=367) | 4.1 | **0.19** | **[0.09, 0.39]** | **0.28** | **[0.13, 0.58]** |
| age at 1st union <20, current age 20-29 (n=11190) | 16.9 | **0.77** | **[0.68, 0.86]** | 1.00 | [0.89, 1.12] |
| age at 1st union <20, current age ≥30 (n=23103) | 31.9 | **1.44** | **[1.30, 1.59]** | **1.40** | **[1.25, 1.58]** |
| age at 1st union 20-29, current age ≥30 (n=14480) | 37.1 | **1.68** | **[1.52, 1.86]** | **1.33** | **[1.18, 1.49]** |
| age at 1st union >=30, current age ≥30 (n=941) | 39.8 | **1.84** | **[1.46, 2.33]** | **1.34** | **[1.05, 1.72]** |

* Among parous women
Results are presented as Prevalence ratios [95% confidence intervals], Bolded results are statistically significant at p-value<0.05
[1] Hypertension defined as SBP≥140 mmHg , DBP≥90 mmHg (average of 2nd and 3rd measurements) or on blood pressure lowering medication or have been told by a doctor/healthcare provider on two separate occasions that they have high blood pressure
[2] Diabetes is defined as NFHS-generated self-report of diabetes mellitus or on glucose lowering medication or have been told by a doctor/healthcare provider on two or more separate occasions that they have high blood glucose levels
[3] Overweight and Obese defined by a body mass index ≥25 $kg/m^2$
[*] Age-adjusted model adjusts for age
[**] Fully-adjusted model adjusts for age, religion, caste, state, place of residence, wealth index, educational attainment, employment status

### References

1. National Family Health Survey (NFHS5), 2019-21: India: Volume I (International Institute for Population Sciences and Inner City Fund) (2021).
